# Circulating tumor DNA precision oncology enables effective and sensitive molecular diagnostics and actionable target detection in pediatric solid tumors - the INFORM experience

**DOI:** 10.64898/2026.01.19.26344140

**Authors:** Kendra K. Maaß, Pitithat Puranachot, Paulina S. Schad, Agnes M.E. Finster, Stefanie Volz, Tom T. Fischer, Barbara C. Jones, Kathrin Schramm, Sophie C. Henneken, Nike Simon, Sophia H. Montigel, Tatjana Wedig, Nathalie Schwarz, Cecilia Zuliani, Petra Fiesel, Christopher Previti, Gnanaprakash Balasubramanian, Florian Iser, Jochen Meyer, Cornelis M. van Tilburg, Till Milde, Olaf Witt, Corinne Rossi, Monika Sparber-Sauer, Stefanie Zimmermann, Thomas Lehrnbecher, Melchior Lauten, Martin Sill, Natalie Jäger, Robert J. Autry, Paul A. Northcott, Felix Sahm, David T.W. Jones, Stefan M. Pfister, Benedikt Brors, Kristian W. Pajtler

**Affiliations:** Faculty of Medicine, Heidelberg University and Heidelberg University Hospital, Department of Pediatric Hematology and Oncology; Hopp Childrens Cancer Center Heidelberg (KiTZ), German Cancer Research Center (DKFZ), German Consortium for Translational Cance; Princess Srisavangavadhana Faculty of Medicine, Chulabhorn Royal Academy, Bangkok, Thailand; German Cancer Research Center (DKFZ), German Consortium for Translational Cancer Research (DKTK), National Center for Tumor Diseases (NCT), Applied Bioinformatics; Heidelberg University Hospital, Department of Pediatric Hematology and Oncology; Hopp Childrens Cancer Center Heidelberg (KiTZ), German Cancer Research Center (DKFZ), German Consortium for Translational Cancer Research (DKTK), National Center for Tumor Di; Hopp Childrens Cancer Center Heidelberg (KiTZ), German Cancer Research Center (DKFZ), German Consortium for Translational Cancer Research (DKTK), National Center for Tumor Diseases (NCT), Heidelberg, Germany, Pediatric Glioma Research; Hopp Childrens Cancer Center Heidelberg (KiTZ), German Cancer Research Center (DKFZ), German Consortium for Translational Cancer Research (DKTK), National Center for Tumor Diseases (NCT), Pediatric Neurooncology, Heidelberg, Germany; Hopp Childrens Cancer Center Heidelberg (KiTZ), German Cancer Research Center (DKFZ), Heidelberg, Germany; Hopp Childrens Cancer Center Heidelberg (KiTZ), German Cancer Research Center (DKFZ) German Cancer Consortium (DKTK), Heidelberg, Germany, Clinical Cooperation Unit Neuropathology; Hopp Childrens Cancer Center Heidelberg (KiTZ), German Cancer Research Center (DKFZ); German Cancer Research Center (DKFZ), Core Facility Omics IT and Data Management, Heidelberg, Germany; Hopp Childrens Cancer Center Heidelberg (KiTZ), German Cancer Research Center (DKFZ), German Consortium for Translational Cancer Research (DKTK), National Center for Tumor Diseases (NCT), Heidelberg, Germany, Pediatric Neurooncology; Faculty of Medicine, Heidelberg University, Heidelberg University Hospital, Heidelberg, Germany, Department of Pediatric Hematology and Oncology; Hopp Childrens Cancer Center Heidelberg (KiTZ), German Cancer Research Center (DKFZ), German Consortium for Translational Cancer Research (DKTK), National Center for Tumor Diseases (NCT), Clinical Cooperation Unit Pediatric Oncology, Heidelberg, Germany; Hopp Childrens Cancer Center Heidelberg (KiTZ), German Cancer Research Center (DKFZ), German Consortium for Translational Cancer Research (DKTK), National Center for Tumor Diseases (NCT), Heidelberg, Germany, Clinical Cooperation Unit Pediatric Oncology; Heidelberg University Hospital, Heidelberg, Germany, Department of Pediatric Hematology and Oncology; Municipal Hospital of the State Capital Stuttgart (gKAoER), Olgahospital, Stuttgart Cancer Center, Center for Pediatric, Adolescent and Womens Medicine, Pediatric Oncology, Hematology and Immunology, Stuttgart, Germany; University Childrens Hospital Tuebi; Goethe University Frankfurt, Frankfurt am Main, Germany, Department of Pediatrics, Division of Hematology, Oncology and Hemostaseology; University of Luebeck, Luebeck, Germany, Pediatric Hematology and Oncology; St. Jude Childrens Research Hospital, Center of Excellence in Neuro-Oncology Sciences (CENOS) and Department of Developmental Neurobiology, Memphis, United States; Faculty of Medicine and Faculty of Biosciences, Heidelberg University; German Cancer Research Center (DKFZ), German Consortium for Translational Cancer Research (DKTK), National Center for Tumor Diseases (NCT), Applied Bioinformatics, Heidelberg, Germany

**Keywords:** Tumor detection, therapy monitoring, pediatric cancer, orthogonal comparison, fragment length, tumor evolution, tumor heterogeneity, precision oncology, tumor board, clinical implementation

## Abstract

Pediatric solid high-risk malignancies mostly lack established molecular biomarkers for early detection, minimal residual disease assessment, or treatment monitoring. Challenges include small patient numbers, limited sample volumes, low tumor mutational burden, and few recurrent alterations. Within the prospective multicenter pediatric precision oncology program INFORM, we collected liquid biopsies from 130 pediatric patients and optimized cfDNA isolation and analysis. Whole-genome, whole-exome, and targeted panel sequencing were performed using liquid biopsy-adapted protocols. Integrating tissue-derived molecular profiles and orthogonal validation revealed that low-coverage whole-genome sequencing reliably detects circulating tumor DNA. An *in silico* ctDNA estimation score, combining fragment length and genome segment alterations, improved sensitivity and specificity to 95%, enabling plasma-based tumor detection in 93% of patients. Whole-exome and panel sequencing effectively identified clinically relevant, potentially druggable molecular targets; however, their utility varied substantially across different tumor entities, underscoring the need for entity-specific considerations in the interpretation and application of these methodologies. In-depth analyses demonstrated liquid biopsy’s potential to track tumor evolution, identifying common tumor ancestors and refining patient stratification. This study advances liquid biopsy methodologies in pediatric oncology and provides a rationale for the molecular alteration-informed selection and implementation of these complementary approaches within molecular tumor boards, with the aim of optimizing clinical management strategies for high-risk pediatric cancer patients.

**Significance:** Systematic liquid biopsy analyses within the pediatric precision oncology INFORM registry enabled a real-world, multicenter comparison of sequencing approaches across high-risk malignancies. By optimizing preanalytical and bioinformatic tools for pediatric settings, we improved plasma-based cancer detection, molecular tumor characterization, and identification of targetable alterations—laying the groundwork for integration into personalized medicine programs and clinical trials.

## Introduction

Children and adolescents with relapsed, progressive, or refractory high-risk malignancies face poor prognoses, prompting the establishment of pediatric precision oncology programs^1^, including pediatric MATCH^2^, MOSCATO-01^3^, MAPPYACTS^4^, the ZERO Childhood Cancer Program^5^, iTHER, SM-PAEDS, PROFYLE, and the INFORM (INdividualized Therapy FOr Relapsed Malignancies in Childhood) registry^6–8^. These programs apply comprehensive molecular profiling of tumor tissue to identify the mechanism-of-action-based treatments. Within INFORM, we validated a predefined molecular target prioritization scheme in a prospective, real-world, multicenter setting^7^. The seven-scale algorithm considers the alteration type and its disease-specific relevance and enables the identification of patient subgroups that clinically benefit from matched targeted treatments by demonstrating extended PFS in patients receiving matched targeted treatment with high-evidence targets^7,8^.

Unlike leukemias, solid pediatric malignancies often lack clonal molecular biomarkers, such as translocations, for continuous disease monitoring. Reliance on radiological imaging delays therapeutic interventions, limiting opportunities for treatment adaptation. To complement tumor analyses, INFORM explored plasma-based liquid biopsies to identify pediatric cancer biomarkers. Liquid biopsies provide a minimally invasive method for real-time tumor monitoring, capturing spatial and temporal genomic changes^9^. While liquid biopsies are increasingly utilized in adult oncology and first FDA-approved diagnostic tests have become available, their implementation in pediatric oncology is challenging due to small patient cohorts, limited sample volumes, and low somatic mutation frequencies^10–14^. The MAPPYACTS trial applied cfDNA whole-exome sequencing (WES) in pediatric solid tumors but achieved a technical success rate in only one-fourth of cases^4^. Additional studies have demonstrated feasibility in specific tumor types but lacked a comprehensive pancancer approach^15,16^. We previously established optimized preanalytical protocols for preserving, isolating, and processing cfDNA across multinational, multi-institutional settings, ensuring reliable genomic analyses with minimal sample input^17^. Our prior work also demonstrated that circulating tumor DNA (ctDNA) serves as a prognostic biomarker for minimal residual disease in medulloblastoma^18^.

To advance liquid biopsy applications in pediatric precision medicine, we implemented a multiplatform sequencing strategy—including whole-genome sequencing (WGS), WES, and targeted panel sequencing—in 130 pediatric patients enrolled in INFORM. Protocols were specifically adapted for small plasma volumes and low cfDNA concentrations. We assessed copy number variations (CNVs), small insertions and deletions (indels), and single-nucleotide variants (SNVs) to enable cancer detection, molecular characterization, and identification of potentially actionable alterations through a tumor type–agnostic approach. To enhance tumor detection sensitivity and specificity, we developed an *in silico* ctDNA estimation score (CES), which was successfully applied across the entire cohort. Liquid biopsies collected during therapy facilitated disease monitoring and enabled early detection of tumor progression—preceding radiological assessments. Notably, direct comparisons revealed that liquid biopsy-based spatial tumor heterogeneity detection provided a more comprehensive molecular profile than traditional bulk tumor analysis, uncovering critical prognostic markers otherwise missed. These findings highlight liquid biopsies as a powerful tool for understanding tumor evolution under treatment pressure, offering novel opportunities for molecularly informed patient stratification and therapy adaptation in pediatric oncology.

## Results

### Characteristics of the INFORM Liquid Biopsy Cohort

The study cohort included 130 pediatric patients enrolled in the INFORM registry between 2015 and 2019, each providing at least one plasma-based liquid biopsy during the current disease episode (Figure 1A, S1A). Molecular profiles of the tumor were generated via the INFORM pipeline, comprising matching tumor and control blood datasets: 130 whole-exome sequencing (WES) pairs, 130 low-coverage whole-genome sequencing (lcWGS) pairs, and 128 tumor methylation array profiles (Figure 1A). The cohort’s demographic, clinical, and sampling characteristics are summarized in Figure S1B and C. Of the cohort, 61 patients had brain tumors, 55 had sarcomas, and 14 had other solid malignancies (Figure 1A). Among 138 initial patients, 8 were excluded due to inclusion criteria failure or tissue quality issues, leaving 130 patients (94%) for analysis via at least one sequencing approach, regardless of plasma volume or cfDNA yield (Figure S1A). cfDNA was extracted from all plasma samples, and the amount of cfDNA, including the first three nucleosomal peaks (160-500 bp, Figure 1B), the total amount of isolated DNA (Figure S1D), and cfDNA purity, expressed as the relative amount of cfDNA per total DNA, were assessed (Figure S1E). Higher cfDNA levels were observed in patients with sarcomas (IQR 22.4–68.0 ng/ml) and other solid tumors (IQR 49.2–79.6.0 ng/ml) compared to brain tumor patients (IQR 0.0–70.5 ng/ml; Figure 1B). To cover genetic alterations such as CNVs, SNVs, and Indels, we performed lcWGS, WES, and targeted panel sequencing. Most samples (26.2 %, n=34) could be analyzed with two methods, while 33.8% (n=44) were even analyzed with three methods (Figure 1C). The final dataset included lcWGS for 133 samples (116 patients), WES for 66 samples (64 patients), and panel sequencing for 74 samples (73 patients) (Figure 1A). Stringent quality criteria defined for all three sequencing platforms were applied, filtering out 4% of analyses (4 lcWGS, 5 WES, 3 panel; Figure S1F). Disease burden potentially affecting liquid biopsy detection is shown in Figure 1D.

**Figure 1.**
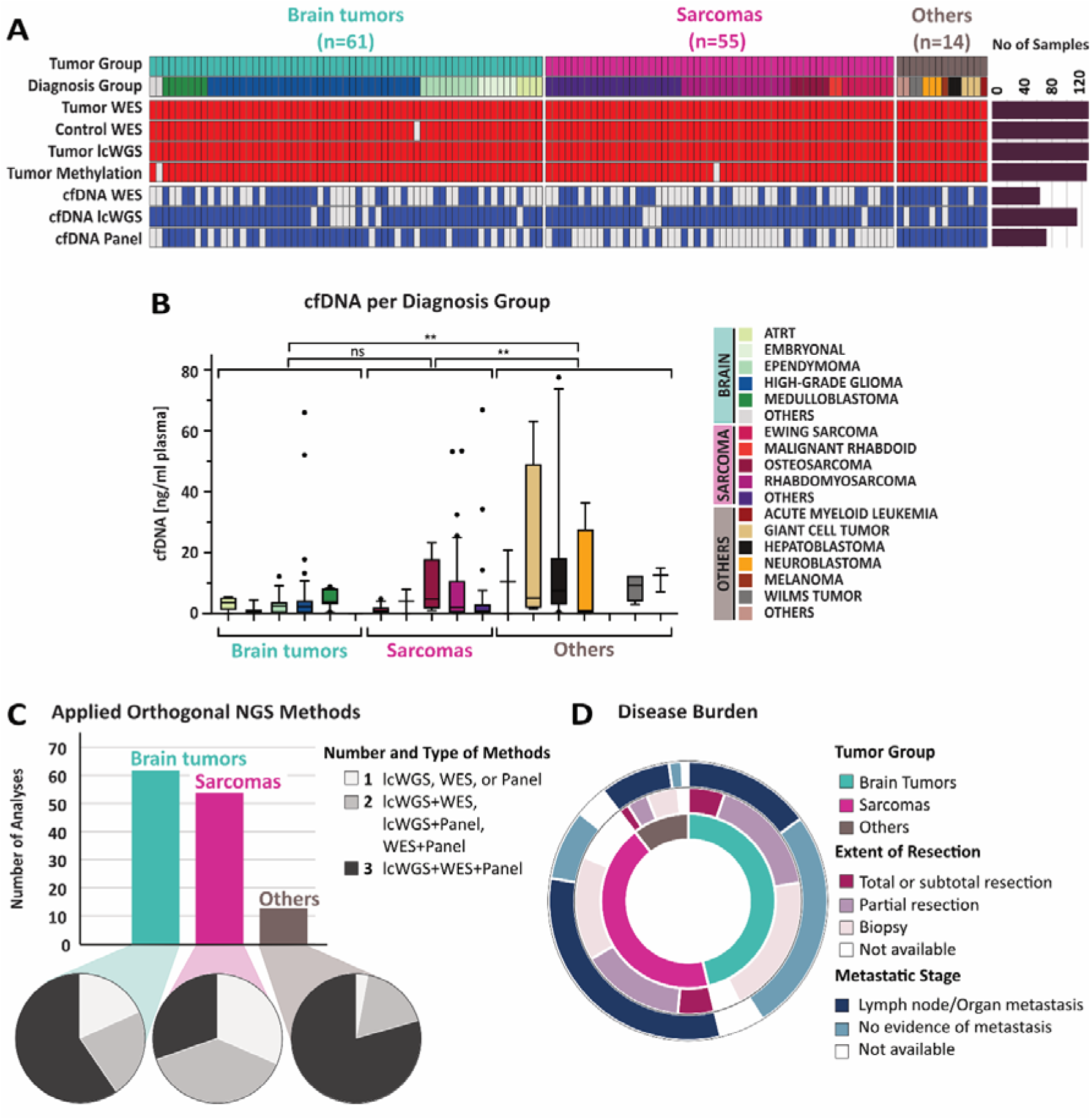
INFORM Liquid Biopsy Cohort Characteristics. **A)** Oncoprint indicating DNA methylation-based diagnosis groups and molecular analyses performed on tumor, blood (control) and plasma-derived cell-free DNA (cfDNA) for INFORM patients with available liquid biopsy material (n=130). **B)** Amounts of cfDNA (ng/ml plasma) isolated from plasma of the indicated diagnosis group. Kruskal-Wallis test (p=0.0027) followed by Dunn’s multiple comparisons test (p>0.9999 for brain tumors vs. sarcomas; p=0.0025 for brain tumors vs. others; p=0.0067 for sarcoma vs. others). **C)** Available orthogonal next generation sequencing (NGS) analyses (low-coverage whole-genome sequencing (lcWGS), whole-exome sequencing (WES) and panel sequencing (panel)) of cfDNA are depicted allowing for two- (n=34) or three-way (n=44) comparisons within respective tumor groups. **D)** Overview of disease burden: tumor group (inner ring), extent of resection (middle ring) and metastatic stage (outer ring).

### The CtDNA Estimation Score (CES) optimizes tumor signal identification using short-fragment data

To overcome volume limitations in pediatric liquid biopsy cohorts, we optimized the workflow, including improved preanalytical protocols for higher cfDNA yields and purity^17^. Adjustments to the WGS library protocol allowed for molecular profiling of plasma samples with cfDNA below the detection limit (50 pg/µl) (Figure S2A1C). Optimized library preparation and tailored amplification cycles allowed detection of plasma-based CNV profiles, even with low cfDNA input (Figure S2D-E). Unique molecular indexing (UMI) strategies reduced amplification bias, increasing average coverage by 8.6%, from 1.27% to 1.38%, for lcWGS (Figure S2F). The fold change in coverage was positively correlated with the duplication rate (Figure S2G).

Using the reported 3% detection limit for the ichorCNA pipeline^19^, we classified patients into high (ichor ctDNA fraction ≥3%) or low ctDNA (ichor ctDNA fraction <3%) (Table S1). Tumor detection rates with ichorCNA were 1.9% for brain tumors, 28.6% for sarcomas, and 20.0% for other malignancies, with 83.3% of samples below the detection limit (Figure 2A, S2H). Recognizing the limitations of ichorCNA, we incorporated cfDNA fragment length as an additional ctDNA indicator. Concordant with previous reports^20^, shorter fragments were found most abundant in high-ctDNA samples (mean length 164.5 bp) compared to low-ctDNA samples (177.4 bp) and healthy controls (179.6 bp, Figure 2B). By using *in silico* size selection for cfDNA fragments smaller than 150 bp (Figure 2C), we markedly increased CNV amplitudes (Figure 2D). The short-to-long-fragment ratio strongly correlated with copy number states, and short fragments were overrepresented in gained and amplified regions (Figure 2E-F). Combining CNV calling with weighted short-fragment data, we developed the ctDNA estimation score (CES), significantly improving tumor detection rates (Method section). CES performance was tested through a titration experiment with ctDNA fractions ranging from 12% to 0.24%. This showed that as ctDNA fractions decreased, the short-fragment ratio increased, correlating well with ctDNA detection (Figure S2I-K). The CES method effectively discriminated between samples with detectable ctDNA and healthy controls, showing a strong correlation with ctDNA fractions (r²=0.85, Figure S2M). In summary, integrating *in silico* size selection of cfDNA fragments with genome segment weighting based on copy number status improved plasma cfDNA detection by approximately 2.5-fold.

**Figure 2.**
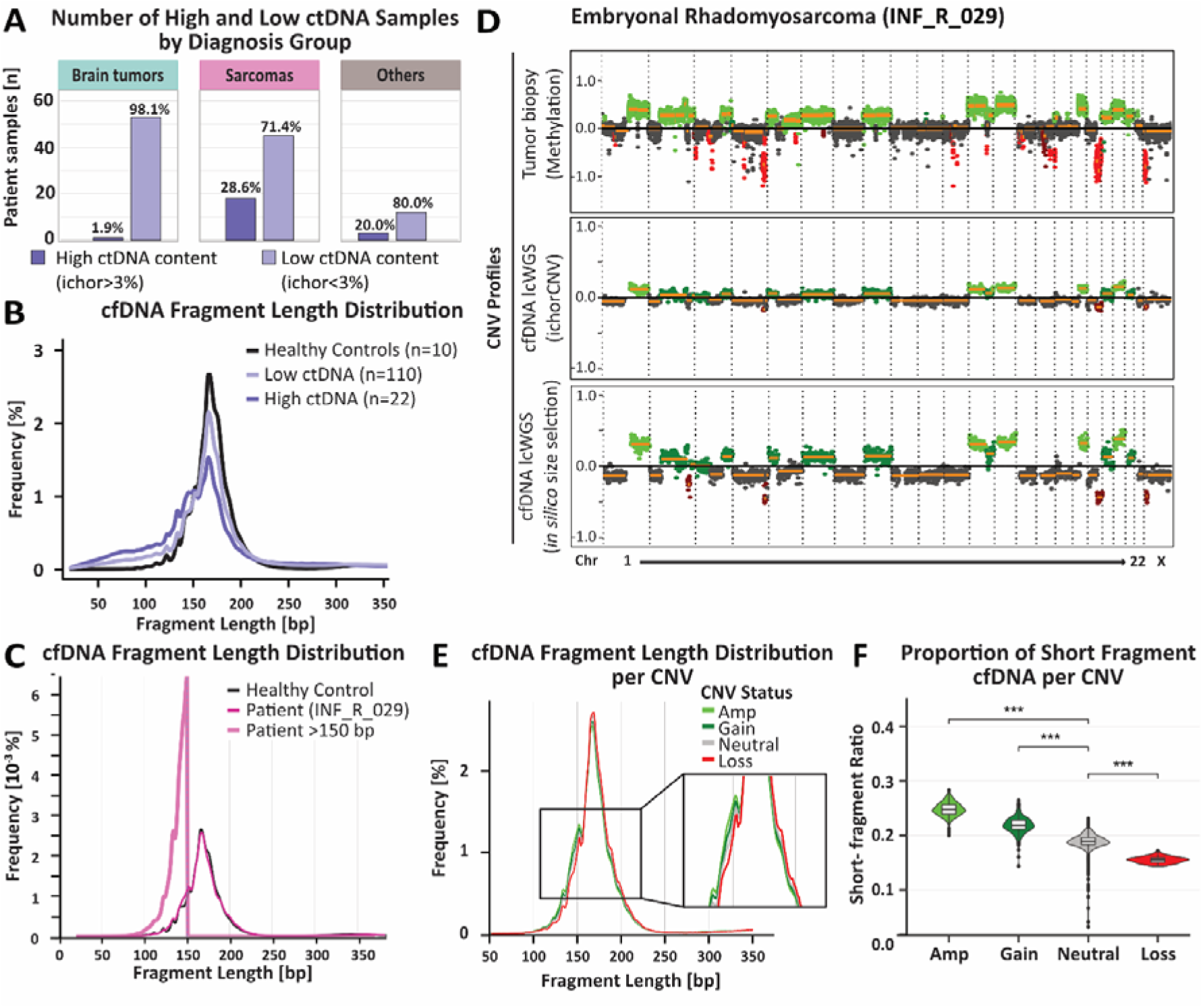
Bioinformatic Refinement of Liquid biopsy-based Low-coverage Whole-Genome Sequencing (lcWGS) to Enhance Tumor Detection from Plasma. **A)** Absolute and relative distribution of brain tumor, sarcoma, and other tumor cell-free DNA (cfDNA) samples with high (ichor>3%, dark purple) or low (ichor<3%, light purple) ctDNA fractions based on ichorCNA. **B)** Average *in silico* fragment-length distribution of plasma-derived cfDNA from patients with high (ichor>3%, dark purple) or low (ichor<3%, light purple) ctDNA fractions plotted against healthy controls (blue). **C)** *In silico*-determined fragment length distribution of cfDNA from a patient tumor (pink, corresponding copy number variation (CNV) plots in **D)** compared to pool of cfDNA of healthy controls (n=10, blue). **D)** Genome-wide CNV profiles of a rhabdomyosarcoma tumor derived from DNA methylation array analysis (upper panel), of a plasma-derived cfDNA sample from the same patient analyzed by lcWGS with the ichorCNA standard pipeline (mid panel) or with *in silico* short fragment (100-150 bp) filtering for improved tumor detection (lower panel). **E)** *In silico*-determined fragment length distribution in relation to the indicated copy-number status. **F)** Violin plot of short-fragment ratio in relation to copy-number aberrations for plasma-derived cfDNA of tumor patient INF_R_029 from C. Compared to CNV neutral regions (grey), the short-fragment ratio was significantly elevated in genomic regions with gains (3N, green) or focal amplification (>3N, light green) and significantly decreased in genomic regions with losses (red). P values were calculated by Wilcoxon rank sum test with *** = p<0.001.

### CES Increases Sensitivity and Specificity of Tumor Detection in the INFORM Liquid Biopsy Cohort

We categorized the liquid biopsy samples into two groups based on ctDNA content: high ctDNA fraction (ichor >3%) and low ctDNA fraction (ichor <3%). We then calculated the ratio of short fragments (100-150 bp) to long fragments (151-250 bp)^20^. This ratio was significantly higher in cancer patients compared to healthy controls. Patients with a high ctDNA fraction had a median ratio of 0.43 (p=0.03), and those with a low ctDNA fraction had a median ratio of 0.22 (p=0.00064), while healthy controls had a median ratio of 0.18 (Figure 3A). The highest ratios were found in sarcoma patients (Figure S3A, S3B). CES significantly improved tumor detection, particularly in brain tumor patients, reaching levels similar to those of solid tumors (Figure 3B-C). The CES demonstrated higher accuracy for tumor detection than ichorCNA, with AUC values of 0.95 versus 0.64 overall (Figure 3D) and 0.93 versus 0.57 for brain tumors (Figure 3E) and 0.96 versus 0.69 for sarcomas (Figure 3F).

**Figure 3.**
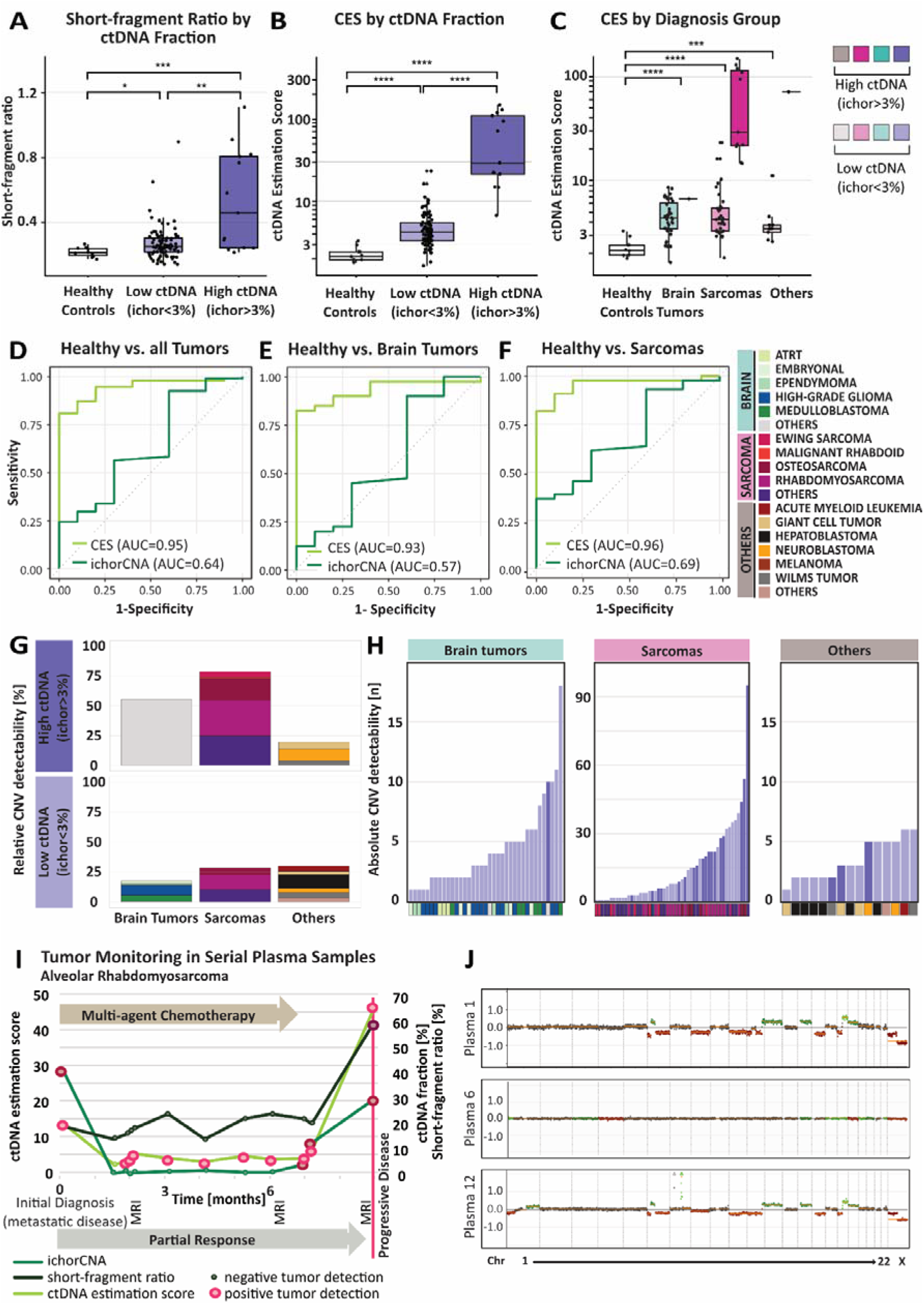
Clinical Validation of Low-Coverage Whole-Genome Sequencing (LcWGS) in a Pediatric Pan-Cancer Liquid Biopsy Cohort. **A)** Short-fragment ratio in liquid biopsy plasma samples of healthy controls, pediatric cancer patients with low (ichor<3%) or high cell-free tumor DNA (ctDNA) fraction (ichor>3%) based on ichorCNA^19^. P-values were calculated by Wilcoxon test with *= p<0.05** = p<0.01, *** = p<0.001. Kruskall-Wallis, p=9.4 × 10^−4^. **B)** CtDNA estimation score (CES) distribution across the same pediatric pan-cancer cohort as in **A)** grouped by ctDNA fraction and compared to healthy controls. P-values were calculated by unpaired t-test with **** = P ≤ 0.0001. Kruskall-Wallis, p=6.2×10 . **C)** CES distribution across the same pediatric pan-cancer cohort as in **A)** and **B)** grouped by both diagnosis group and ichorCNA ctDNA fraction. D-F) Receiver Operating Characteristic (ROC) curves illustrating increased sensitivity and specificity of CES to ichorCNA ctDNA fraction analyses for **D)** healthy controls versus pan-cancer cohort (Area under the curve (AUC) CES=0.95, ichorCNA=0.64;); **E)** versus brain tumor patients (AUC CES=0.93, ichorCNA=0.57), and **F)** versus sarcoma patients (AUC CES=0.96, ichorCNA =0.69). G-H) Bar plots showing **G)** relative and **H)** absolute numbers of detected tumor tissue CNVs in plasma cfDNA per diagnosis group. Plasma samples with ctDNA fraction ichor ctDNA fraction<3% are labeled in light purple, plasma samples with ctDNA fraction ichor ctDNA fraction >3% are labeled in dark purple. **I)** IchorCNA ctDNA fraction, short-fragment ratio and CES of cfDNA from an alveolar rhabdomyosarcoma patient collected over 7 months. Red circles indicate positive tumor detection, highlighting the detection of molecular residual disease (MRD) 2 months prior to magnetic resonance imaging (MRI) diagnosis of progressive disease. **J)** Genome-wide CNV profiles of plasma cfDNA from rhabdomyosarcoma patient collected serially during therapy course.

Expanding the analysis beyond tumor detection, we assessed the concordance between plasma cfDNA and matched tumor CNV profiles using lcWGS data, which includes clinically relevant CNVs from the INFORM list of actionable targets. Among brain tumors, the only high-ctDNA sample came from a medulloblastoma patient with metastatic spread (M4) and residual tumor mass (R4), showing strong agreement between cfDNA and tumor-derived CNV profiles, with 55% (10/18) of tumor CNVs detected (Figure 3G (upper left) and 3H (dark purple)). In low-ctDNA brain tumors, high-grade gliomas and medulloblastomas had the highest CNV detectability (Figure 3G (lower left) and 3H (light purple)). Sarcomas in the high-ctDNA group had the highest CNV detection rate (79%; Figure 3G, upper middle), with osteosarcoma, rhabdomyosarcoma, and other sarcomas showing similar detection rates across high (74%) and low (28%) ctDNA groups (Figure 3G, middle, Figure 3H). Among other tumors, neuroblastomas dominated successful CNV detection in high-ctDNA cases (Figure 3G, upper right), while low-ctDNA tumors had comparable CNV detection rates (30%) to sarcomas (Figure 3G, lower right). Notably, sarcoma plasma CNV profiles in high-ctDNA cases closely mirrored tumor tissue alterations, reinforcing their clinical relevance (Figure S3H).

Finally, we assessed longitudinal tumor monitoring using serial plasma samples (n=12) from a patient with metastatic alveolar rhabdomyosarcoma (Figure 3I). Following an initial partial response (PR) to multi-agent chemotherapy, residual tumor was monitored through routine radiologic imaging. While ctDNA fractions and short-to-long fragment ratios remained below detection limits (Figure 3I, ctDNA fraction—dark green, short-to-long ratio—olive), the CES score effectively captured the remaining tumor burden (CES—light green). Seven months post-diagnosis, both CES and short-to-long fragment ratios signaled progressive disease (PD), detecting it two months earlier than routine imaging performed at three-month intervals (Figure 3I). To further validate the clinical relevance of the CES score, we analyzed a previously published longitudinal cfDNA dataset from a patient with Ewing Sarcoma^21^. A total of six cfDNA samples were collected across the clinical timeline, including diagnosis, treatment, and relapse phases. As depicted in the time resolved analyses tumor-derived signal in cfDNA increased substantially during relapse periods with ichorCNA-estimated ctDNA fraction rising from 0.09 at diagnosis to 0.55 at first relapse, and remaining elevated at 0.42 during the second relapse (Figure S4A). In parallel, the CES score showed a concordant pattern with disease progression remaining low at diagnosis, peaking at first relapse, transiently declined following treatment, and rose again at secondary relapse (Figure S4B, S4C). Notably, it remained elevated prior to the patient’s death on Day 654.

These findings underscore that the CES score mirrors clinical tumor burden and aligns with established measures such as ichorCNA ctDNA fraction. This demonstrates the potential of CES as sensitive, non-invasive biomarker for real-time monitoring of therapy response in the future in pediatric cancers with low mutational burden.

### Whole-Exome and Targeted Panel Sequencing for Detecting Druggable Mutations in cfDNA

Beyond CNV-based tumor detection, liquid biopsies in personalized oncology require mutation analysis to assess target abundance and detect resistance early. We applied WES and targeted panel sequencing to cfDNA samples with sufficient material, enabling deeper coverage and analysis (Figure S5A and G). WES-based detection of somatic SNVs was assessed in 28 brain tumors, 26 sarcomas, and 12 other pediatric cancers, focusing on 367 clinically relevant genes (Table S2, Figure 4, Figure S5). Average detection rates varied across tumor types (brain tumors: 11.3%, sarcomas: 32%, others: 27%; Figure 4A-C). Patient-specific alterations are shown for each diagnosis group as both absolute counts and relative proportions of tumor-informed cfDNA SNVs, emphasizing the detection rates of clinically relevant mutations (Figure S5B-D). Among sarcomas, 9 cfDNA samples had druggable alterations, 67% matching tumor tissue results, while 33% contained additional cfDNA-exclusive SNVs (Figure 4B, S5C).

**Figure 4.**
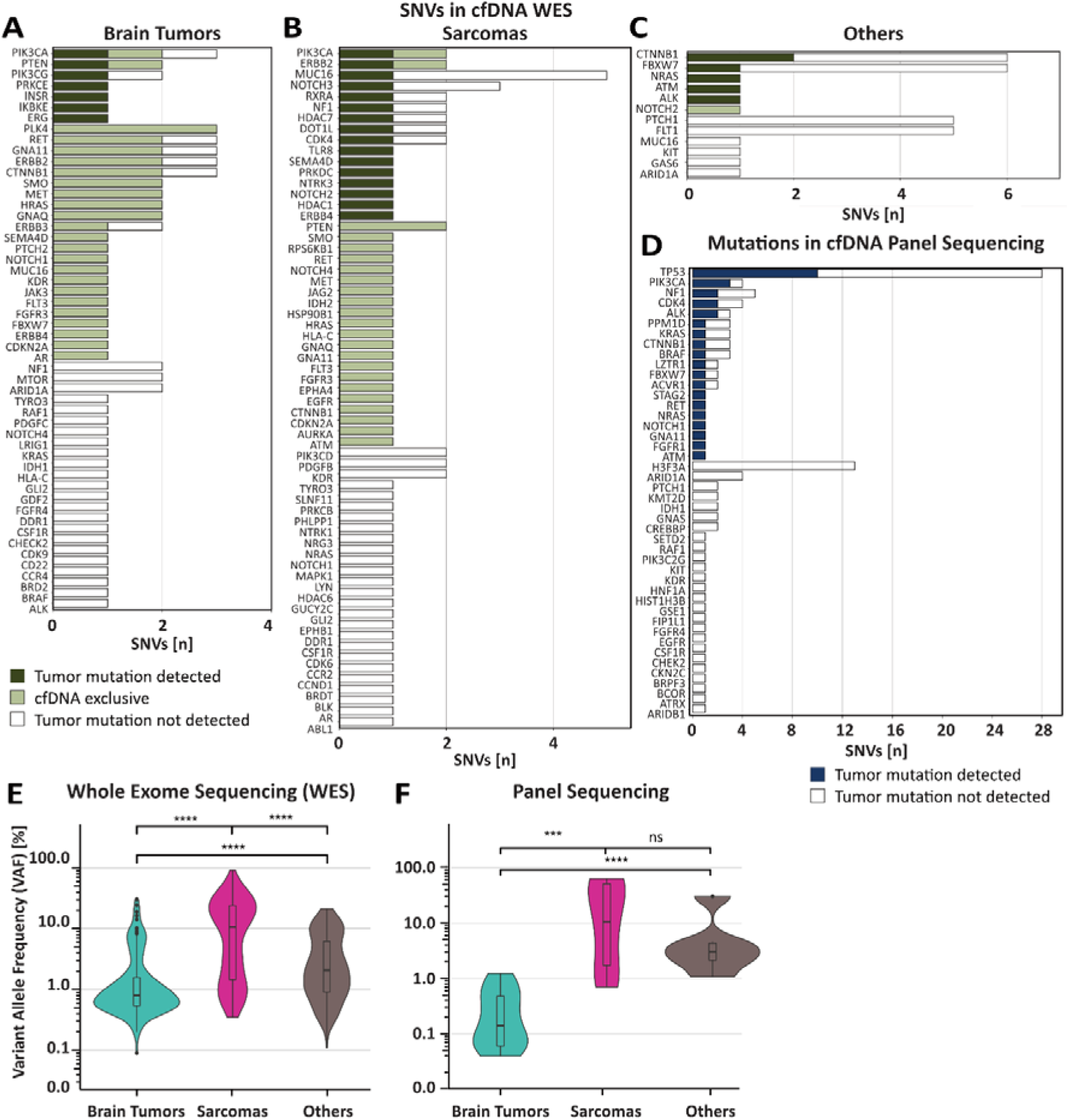
Whole-Exome Sequencing (WES) and Targeted Panel Sequencing Approaches Complemented Low-Coverage Whole-Genome Sequencing (LcWGS) through Detection of Single Nucleotide Variants (SNVs) **A)** SNVs detected in cell-free DNA (cfDNA) of **A)** brain tumor, **B)** sarcoma, and **C)** other tumor patients by WES and **D)** panel sequencing analysis. Dark green/blue – somatic tumor SNV detected in cfDNA, light green – cfDNA exclusive SNV, white – somatic tumor SNV not detected in plasma. **E-F)** Variant allele frequency (VAF) of SNVs detected in cfDNA for indicated diagnostic groups **E)** by WES and **F)** by panel sequencing.

For panel sequencing, UMI barcoding increased read recovery threefold (median depth: 328.5 to 820.5) (Figure S5E), enhancing SNV detection at lower variant allele frequencies (VAF) (Figure S5F-G). The most recurrent SNVs in this cohort were in TP53, PIK3CA, NF1, CDK4, and ALK (Figure 4D). Reducing the panel to 130 genes^22^ improved relative detection rates across all groups (25-100%, Figure S5H1J), with tumor-informed SNVs found in 21% of brain tumors (Figure S5H), 71% of sarcomas (Figure S5I), and 40% of other tumors (Figure S5J). Panel sequencing had greater sensitivity in brain tumors, detecting SNVs with a minimum VAF of 0.04% (vs. 0.09% in WES, (Figure 4E-F, S5), but did not improve detection in sarcomas (median VAF WES 14.69% versus panel 24.58%) or other tumors (median VAF WES 4.19% versus panel 7.34). This highlights panel sequencing’s utility for low-VAF SNVs, particularly in brain tumors for which the panel was designed.

### Performance Comparison of cfDNA Sequencing Approaches

We compared lcWGS, WES, and panel sequencing for detecting tumor-derived alterations in plasma-based liquid biopsies, using tumor tissue alterations detected by lcWGS (CNVs) or WES (SNVs) as ground truth (Figure 5). For CNV detection, lcWGS showed a higher detection rate (≥1 CNV detected, 88% light blue) compared to WES (25%, dark green, Figure 5A), with sarcomas exhibiting the highest detection rates (lcWGS 90%, WES 52%), followed by brain tumors (lcWGS 79%, WES 8%) and other pediatric cancers (lcWGS 100%, WES 14%). The number of tumor CNVs (median 18, maximum 291) did not affect detection rates. In terms of SNVs, WES identified more tumor-specific alterations than lcWGS (WES: 84%, lcWGS: 21%), with insertions and deletions (Indels) detected in 42% of tumors by WES and 6% by lcWGS (Figure S6A, B). The sequencing depth (130x for WES vs. 3x for lcWGS), coverage uniformity (with missing reads for lcWGS indicated), and input cfDNA amounts (mean 41 ng for WES vs. mean 139 pg for lcWGS) led to superior detection of somatic SNVs in liquid biopsies by WES (965/4081) compared to lcWGS (52/4081, Figure S6A).

**Figure 5.**
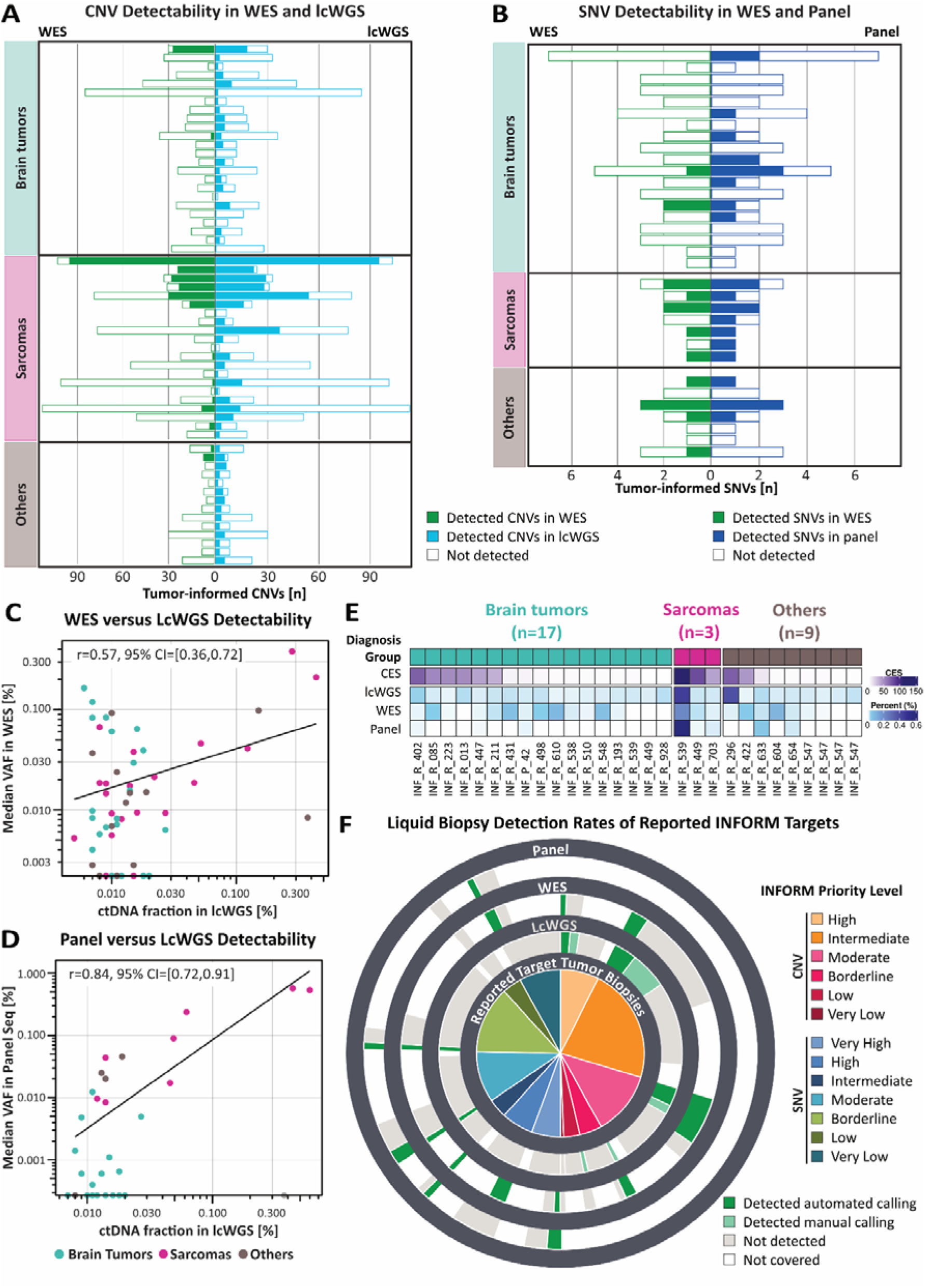
Comparison of Orthogonal NGS Approaches on cell-free DNA (cfDNA) Liquid Biopsies. **A)** Orthogonal comparison of copy number variation (CNV) detectability in liquid biopsy whole-exome sequencing (WES) (left bars) versus low-coverage whole-genome sequencing (lcWGS) (right bars) in matched patient plasma samples. Green – tumor CNVs detected in cfDNA WES, light blue – tumor CNVs detected in cfDNA lcWGS, white – tumor CNVs not detected by the respective method. **B)** Orthogonal comparison of single nucleotide variant (SNV) detectability in liquid biopsies in WES (left bars) versus panel sequencing data (right bars) in matched patient plasma samples. Green – common SNVs in tumor and cfDNA detected in cfDNA WES, dark blue – common SNVs in tumor and cfDNA detected in cfDNA lcWGS, white – tumor SNVs not detected by the respective method. **C-D)** Correlation plots of median variant allele frequencies (VAF) of detected tumor-informed mutations in **C)** WES and **D)** panel data with lcWGS-derived cell-free tumor DNA (ctDNA) fractions of matched cfDNA samples. **E)** Direct comparison of liquid biopsy-based tumor detection ability by plotting CES, ichorCNA ctDNA fraction (%) in lcWGS, and median VAF (%) of tumor-informed mutations in WES or panel sequencing in matched samples of 29 patients. Diagnosis groups are indicated. Discrepancy between the three methods was observed for brain tumor patient 11, 15 and 16 (INF_R_431, INF_R_610, INF_R_548) and other tumor patients (INF_R_296, INF_R_604). **F)** Sunburst plot summarizing cfDNA-based detection of targets (CNVs/SNVs) that were reported for corresponding tumors in the INFORM molecular tumor board. Target priority levels are indicated in the inner circle. Dark green – alteration called with automated bioinformatics pipeline, light green – alterations called through visual inspection, red – alteration not detected despite being covered in data of the indicated method, white – alteration not covered in data of the indicated method.

When comparing panel sequencing to WES, the panel detected tumor-informed SNVs in 55% of patients, outperforming WES, which detected SNVs in 33% (Figure 5B). The increase in SNV detection was particularly notable in brain tumors (Panel: 42%, WES: 11%) and sarcomas (Panel: 100%, WES: 71%). For other malignancies, WES slightly outperformed the panel (Panel: 43%, WES: 57%, Figure 5B). We also compared ctDNA fraction and variant allele frequency (VAF) between WES and panel sequencing (Figure 5 C-D, Figure S6C-D). The WES median VAF exhibited a weak correlation with the lcWGS-based ctDNA fraction (r=0.57, Figure 5C), likely due to subclonal SNVs. This correlation improved (r=0.67) when using the maximal VAF instead (Figure S6C). Panel sequencing had a stronger correlation with lcWGS ctDNA fraction (r=0.84 for median VAF and r=0.83 for maximal VAF Figure 5D, S6D). Sample-matched analyses (n=29) revealed good concordance across sequencing methods (Figure 5E). lcWGS was superior in identifying high-priority CNVs, while panel sequencing excelled at detecting SNVs with very high priority (Figure 5F, Figure S6E). The selection of sequencing approach should be tailored to the specific diagnosis, liquid biopsy purpose (such as minimal residual disease vs. target detection), mutation types, and cfDNA yield.

### Liquid Biopsy-Based Detection of Temporal Heterogeneity

Liquid biopsies can reflect genetic alterations across different stages of a tumor, aiding therapeutic target identification and disease monitoring. By analyzing serial liquid biopsies from primary diagnosis, relapse, and metastasis, we assessed temporal tumor evolution (Figure 6) and spatial heterogeneity (Figure 7). Patient INF_R_425, diagnosed with osteosarcoma of the right humerus, underwent surgery and chemotherapy, followed by metastases and spinal lesions. The occurrence of a spinal lesion triggered enrollment in the INFORM registry at month 52 after diagnosis, with a liquid biopsy plasma sample being available alongside material from the primary tumor and a lung metastasis (41 months post-diagnosis) for parallel molecular analysis (Figure 6A). lcWGS CNV analysis of the plasma sample revealed chromosomal instability, with VEGFA amplification and CDKN2A/B deletion present in the primary tumor and spinal lesion, but only VEGFA amplification in the lung metastasis (Figure 6B, Figure S7A-B). Comparison of structural rearrangements in Circos plots highlighted distinct genomic instability patterns at each sampling site (Figure 6C, S7C). To investigate tumor evolution, we inferred clonal hierarchies using VAF-based SNV clustering from all tumor tissues and plasma (Figure S7D). The primary tumor had four subclonal lineages, three of which were detectable in the plasma (Figure 6D). Subclone 1 (54%) was stable across metastasis, and predominated in the cfDNA (77%), while subclone 2 (46%) became extinct. Metastatic subclones 3 and 4 from the lung (39%, months 41) and spinal metastases (31%, months 52) were both detected in plasma (15% and 8%, respectively). Subclone 1 remained predominant, with cfDNA composition closely resembling the spinal lesion (Figure 6D).

**Figure 6.**
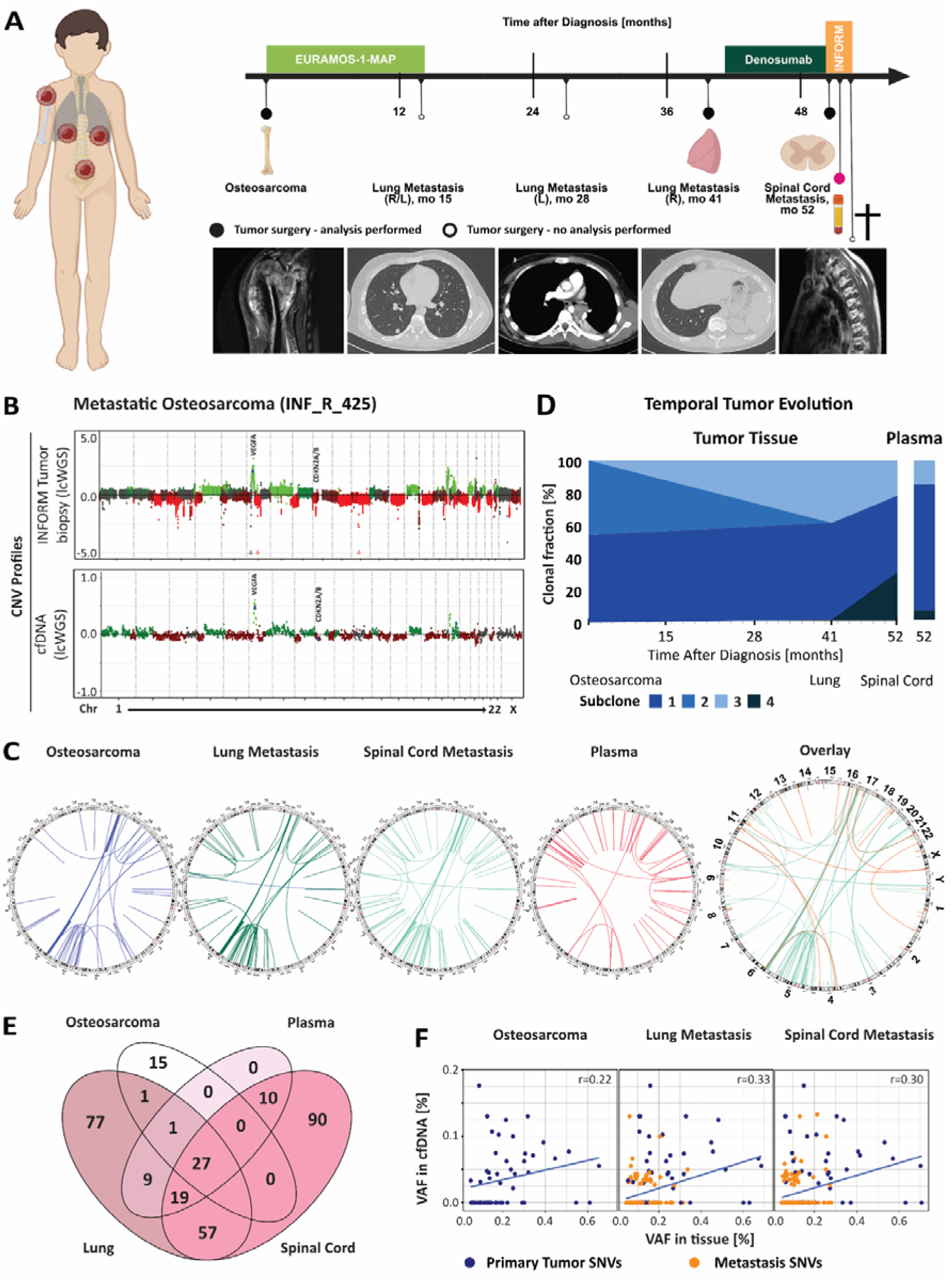
Detection of Temporal Tumor Heterogeneity in Liquid Biopsy Samples. **A)** Graphical summary of tumor sites diagnosed over time for an individual INFORM patient with metastatic osteosarcoma. Black dots indicate tumor episodes with available molecular tumor data, black circles indicate tumor episodes without available molecular tumor data. **B)** WGS-derived CNV profiles of a tumor tissue biopsy (upper) and of a plasma-derived cfDNA sample obtained following surgery analyzed with *in silico* short fragment filtering for improved tumor detection (lower). Both VEGFA (chromosome 6p) and CDKN2A/B (chromosome 9p) amplifications that were reported by the molecular INFORM tumor board for the tissue biopsy (52 months after diagnosis) were also detected in a simultaneously obtained plasma sample. **C)** Circos plots of structural variants per indicated site and as overlay plot of the primary osteosarcoma (blue), a lung metastasis (41 months after diagnosis, green), a spinal cord metastasis (52 months after diagnosis, turquois) and plasma cfDNA (52 months after diagnosis, red). **D)** Temporal evolution analysis of tumor tissues biopsies from indicated time points based on WES-derived VAF of SNVs. Subclonal assignment to the cfDNA sample revealed a major subclone already present at primary diagnosis and two minor subclones originating from subsequent lung and spinal cord metastatic sites. **E)** Venn diagram of somatic SNVs revealing that the liquid biopsy analysis reflects clonal contribution from the different tumor sites. **F)** Evolution of tumor SNVs and their respective VAF in tumor tissue (primary/metastasis, WES) and plasma-derived cfDNA (lcWGS). Correlation plot of VAF in cfDNA and tissue of indicated tumor sites. Dark blue indicates SNVs detected in the primary osteosarcoma, orange indicates SNVs that evolved over time (*r=0.22 (Osteosarcoma), r=0.33 (Lung Metastasis), r=0.33 (Spinal Cord Metastasis*).

**Figure 7.**
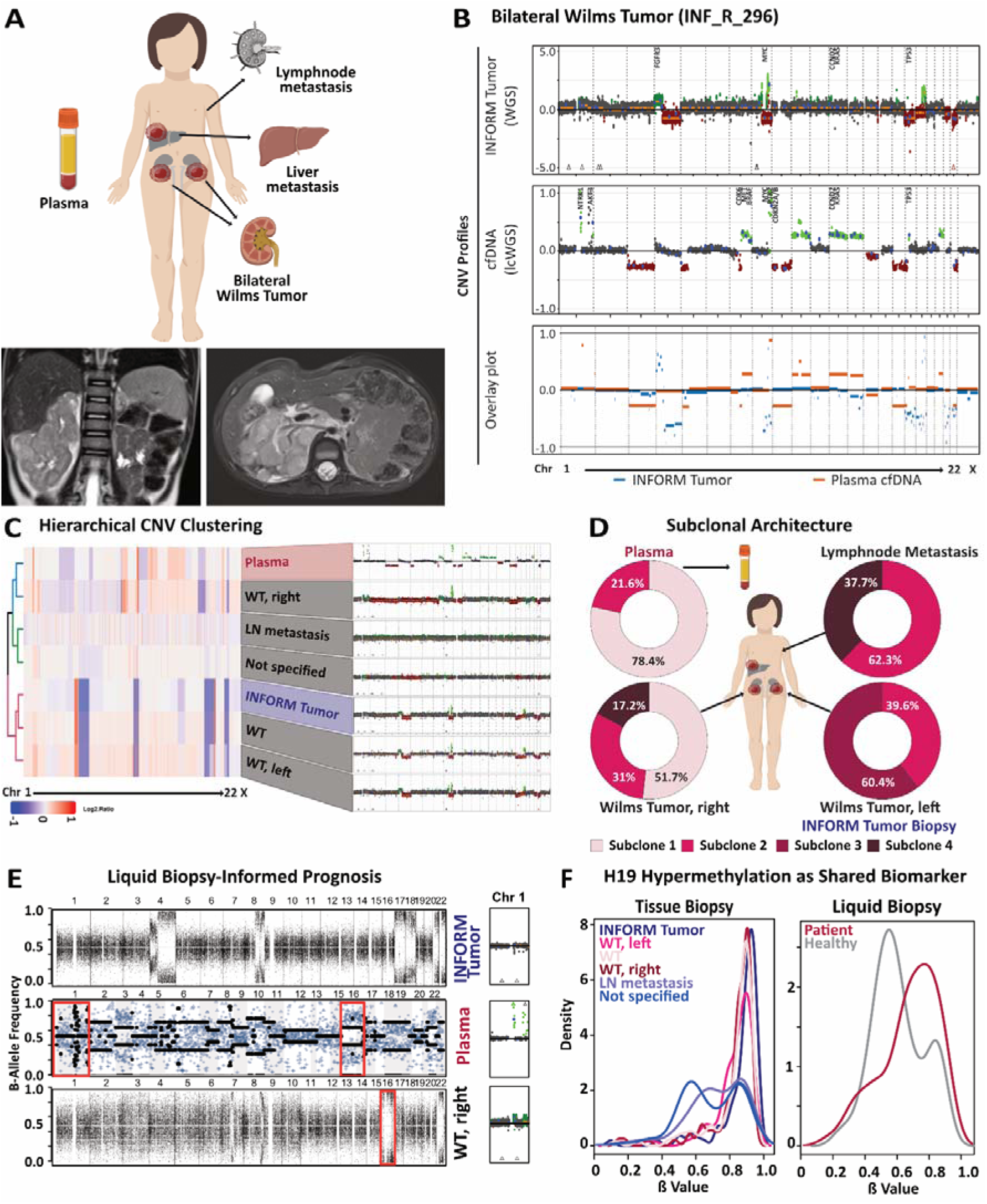
Liquid biopsy-based Detection of Spatial Tumor Heterogeneity. **A)** Graphical summary of tumor diagnosis at different locations for an individual INFORM patient with bilateral, metastatic Wilms tumor (WT). MRI of the indicated patient highlighting the liver metastasis. **B)** Genome-wide CNV profiles derived from WGS of tumor tissue biopsy (upper), cfDNA lcWGS (middle) and overlay plot of tumor (blue) and plasma (orange, lower). Colors of the CNV profile represent neutral CNV state (grey), deletions (red), gains (3N, green) and focal amplifications (>3N, light green). **C)** Unsupervised hierarchical clustering of CNVs by Pearson’s correlation identified the WT from the right site to be the predominant plasma clone. There is a discordance of the cfDNA and the WT sample analyzed for the INFORM molecular tumor board. Corresponding CNV profiles are highlighted on the right. **D)** Subclonal architecture within tumors from indicated sites and plasma cfDNA based on WES-derived VAF of SNVs. Subclonal assignment to the cfDNA sample revealed a major subclone (Subclone 1, 78.4%) unique for WT from the right site (Subclone 1, 51.7%), which is not present in the WT sample analyzed for the INFORM molecular tumor board (biopsy from the left side). **E)** B-allele frequency plot showed no loss-of-heterozygosity (LOH) of chromosomes 1q or 16p in the INFORM tumor tissue analysis. In contrast, the plasma sample presented partial LOH on chromosome 1 and a distinct LOH on the entire chromosome 16 (highlighted in red), stratifying patient a poor prognosis. **F)** ß-Value density plot of the H19 locus revealed H19 hypermethylation as shared early oncogenic hit of all investigated tumor tissue biopsies. In plasma liquid biopsies, H19 methylation patterns allowed a clear discrimination of the WT patient from healthy controls.

We also assessed the impact of sequencing method and coverage on SNV detection and subclonal composition in plasma cfDNA using low (1x) and medium (16x) coverage WGS, WES, and panel sequencing (Figure S7E-H). lcWGS detected shared and unique CNVs, and WGS/WES showed positive correlation between SNV amounts and genomic coverage (Figure S7E-G), while the panel covered only a single SNV (Figure S7H). We examined mutation representation over time across tumor sites: primary osteosarcoma (64%, diagnosis), lung metastasis (23%, month 41), and spinal cord metastasis (28%, month 52) in cfDNA (Figure 6E). Plasma-based detection of unique SNVs further supported the resemblance of cfDNA to the spinal lesion, with 10 unique SNVs from the spinal metastasis, 1 from the lung, and none from the primary tumor (Figure 6E). VAF-based correlation analyses showed that as the disease progressed, the VAF correlation between liquid biopsy and tumor sites increased, indicating that plasma-based liquid biopsy effectively reflected the evolving subclonal architecture (Figure 6F, Figure S7F). These results suggest that temporal tumor evolution can be monitored via cfDNA analysis.

### Liquid Biopsy Captures Spatial Tumor Heterogeneity, Enabling Location-Independent Molecular Profiling

Performance comparisons revealed high plasma tumor content but low concordance with tissue SNVs and CNVs for Patient 25 (INF_R_296), diagnosed with metastatic bilateral Wilms tumor progressing under first-line SIOP 2001 GPOH therapy (HR, Stage V, Figure 7A). At INFORM enrollment, metastases were present in the liver, lymph nodes, and abdominal wall (Figure 7A). The INFORM pipeline detected MYC amplification, TP53 mutation, and KDM1A overexpression in a Wilms tumor sample (=INFORM tumor) without a specified location (Figure 7B, S8A). For the liquid biopsy at enrollment, ichorCNA reported 37.4% ctDNA with distinct alterations, including focal amplifications on chromosome 1 and whole chromosome gains of 10, 12, and 13 (Figure 7B). CNV analysis showed overlapping but also unique aberrations between tumor and plasma (Figure 7B). Genotyping confirmed sample identity (Figure S8B). Tracing cfDNA origins, WGS and methylation analysis of primary tumors from both kidneys, an unspecified site, and lymph node metastasis revealed that the INFORM tumor corresponded to the left Wilms tumor, while the right tumor was the predominant cfDNA source (Figure 7C). Correlation analysis showed a Pearson coefficient of 0.7 between cfDNA and the right tumor vs. 0.04 for the left tumor (Figure S8C). Methylation confirmed these findings (Figure S8D-E).

SNV-based phylogenetic analysis identified two dominant subclones in cfDNA (78% and 22%), with the latter shared across all tumor sites (Figure 7D). However, the most prominent INFORM tumor subclone was absent from cfDNA, highlighting site-specific divergence (Figure 7D, S8E). Notably, three gene alterations were cfDNA-exclusive, while two were unique to the INFORM tumor (Table S3). These findings underscore the need for further validation of liquid biopsy-detected targets for clinical actionability.

Furthermore, we investigated prognostic Wilms tumor biomarkers with potential relevance for therapy intensification^23,24^. Loss of heterozygosity (LOH) on 1p/16q was detected in plasma but not in the INFORM tumor, confirming its right Wilms tumor origin (Figure 7E, left). The prognostically relevant 1q gain appeared in cfDNA as a focal amplification (up to 10 copies) and was traced to the right tumor, where it spanned a broader region (Figure 7E, right). To identify a shared Wilms tumor biomarker, we analyzed H19 locus methylation across tumor sites^25^. All tumors with recognizable content showed clear H19 hypermethylation, while the lymph node metastasis and a low-content sample exhibited an indistinct ß-value distribution (Figure 7F, left). Notably, H19 hypermethylation was also detected in cfDNA, distinguishing tumors from normal controls (Figure 7F, right).

These findings illustrate how liquid biopsies capture regionalized risk factors undetected by single-tumor analysis, potentially informing (neo)adjuvant strategies. H19 hypermethylation may serve as a universal Wilms tumor biomarker, warranting further investigation for clinical implementation.

## Discussion

The INFORM registry has demonstrated the feasibility of comprehensive molecular profiling for pediatric tumors in a real-world, multi-institutional setting^7^. However, high-level evidence targets for matched therapies that improve clinical outcomes remain limited (∼10%) and cannot be serially monitored via invasive biopsies^7,8^. To address this, pediatric precision oncology programs are integrating drug sensitivity profiling^26^, proteomics, and liquid biopsy to capture spatial and temporal tumor heterogeneity. Here, we present a comparative plasma-based DNA sequencing approach for tumor detection, molecular profiling, and target identification in high-risk pediatric brain and solid tumors. In line with previous studies, cfDNA levels were higher in patients with certain solid tumors, such as neuroblastoma^4^. Although brain tumors represent the largest diagnostic group and plasma cfDNA is typically considered an unreliable liquid biopsy source, our combined analytical approach successfully detected tumors in the majority of patients. With a 93% detection rate, our method demonstrates the potential of liquid biopsy for molecular residual tumor assessment, even in challenging cases. Our study confirmed WES superiority for target detection, consistent with the MAPPYACTS study^4^. However, non-standardized sample collection across clinical sites posed challenges, with some cfDNA samples showing high genomic DNA contamination. To address this, we applied an optimized preanalytical workflow, including UMI integration, low-input library construction, and short DNA fragment enrichment to ensure data quality^17^.

Liquid biopsy-based molecular residual disease (MRD) detection strongly predicts recurrence in adult cancers and may guide treatment decisions^27–33^. However, its clinical utility in pediatric oncology remains under investigation. Our study, focused on high-risk patients, achieved a 95% tumor detection rate by integrating *in silico* DNA size selection and segmental copy number analysis within our CES algorithm. Notably, CES identified tumor progression months before clinical detection, similar to adult liquid biopsy studies^33,34^. Further evaluation is needed, particularly in first-line settings with lower ctDNA burden, and for its prognostic and response-monitoring utility^35^.

Pediatric cancers exhibit genetic heterogeneity between primary and metastatic sites^36–39^. In exemplary cases, plasma-exclusive findings were mapped to previously unsampled lesions, highlighting the power of minimally invasive technologies to capture clinically relevant subclones for targeted therapy and risk stratification. Our data support an integrated liquid biopsy workflow combining cfDNA screening via lcWGS with alteration-adapted analysis, complementing molecular tumor profiling within precision oncology programs like INFORM. The selection of sequencing approaches, data interpretation, and quality measures necessitates clinical decision support tools for streamlined translation into clinical practice.

Our pancancer liquid biopsy study demonstrated the effectiveness of three NGS technologies in an orthogonal comparison, achieving a 92% technical success rate despite small sample volumes and low genetic redundancy. Our LiQuiTect pipeline is now integrated into INFORM as an additional molecular layer. Ongoing prospective applications within early-phase and therapy optimization trials^7,40^ will further define the clinical impact of liquid biopsy in pediatric oncology.

## Materials and methods

### Study design, eligibility and participants

The INFORM registry is a prospective, noninterventional, multicenter, multinational, feasibility registry that collects clinical, functional (biological), and molecular data. After a pilot phase^6^, the registry opened on January 21st, 2015. The study was registered at the German Clinical Trial Register with the number DRKS00007623. Ethics committee approval for conducting the study, the use of consent forms and scientific evaluation of the data were obtained from Heidelberg University’s review board (S-502/2013; S-795/2020). Eligible patients included children, adolescents and young adults with refractory, relapsed or progressive oncological disease after initial treatment according to the GPOH protocol, as well as specific primary indications. The detailed inclusion criteria are outlined in Worst *et al*. 2016^6^. The tumor samples and matched germline controls from each patient were subjected to whole-exome sequencing (WES), low-coverage whole-genome sequencing (lcWGS), RNA sequencing (tumor only) and methylation array (450K, 850K or EPIC array, tumor only) as previously described^6^. All patients whose plasma samples were available (01/2015-08/2019) were included in this study. Blood samples were collected via standard venipuncture, processed at the respective treatment center and shipped together with tumor and germline material. Blood samples were collected before and after surgery. All the plasma samples were stored at -80°C until further processing. For healthy controls, peripheral blood samples were collected from 10 healthy individuals aged between 24 and 48 years by standard venipuncture in EDTA tubes and centrifuged at 4°C for 10 min at 1900 × g.

### cfDNA isolation from plasma samples

Supernatants from preprocessed plasma samples were thawed on ice, and cfDNA was isolated via the NucleoSnap cfDNA Kit (Machery Nagel, Düren, Germany) according to the manufacturer’s manual. Elution was performed with 50 µl of nuclease-free water (Thermo Fisher Scientific, Waltham MA, USA). The extracted cfDNA was quantified with a Qubit dsDNA HS Assay Kit (Thermo Fisher Scientific, Waltham, MA, USA), and the fragment size distribution was assessed with a Bioanalyzer High Sensitivity DNA Kit (Agilent, Santa Clara, CA, USA). All cfDNA samples were stored at −80°C until further analysis.

### Low-coverage whole-genome sequencing of cfDNA

WGS libraries were prepared with an Accel-NGS 2S Hyb DNA Library Kit (Swift Biosciences, Ann Arbor, MI, USA) according to the manufacturer’s protocol. cfDNA was used without fragmentation, and when available, the input was adjusted to 100 pg per library. Libraries were ligated with single-indexing adapters and unique molecular identifiers (UMIs; Swift Biosciences, Ann Arbor, MI, USA). The number of amplification cycles was adjusted from 12 to 16 cycles depending on the input yield. No-template and water controls were included in each experiment. Libraries were quantified with a Qubit dsDNA HS Assay Kit (Thermo Fisher Scientific, Waltham, MA, USA), and the fragment size distribution was assessed with a Bioanalyzer High Sensitivity DNA Kit (Agilent, Santa Clara, CA, USA). Libraries were multiplexed in equimolar amounts and sequenced at 100 bp paired ends on a NovaSeq 6000 S4 flow cell (Illumina, San Diego, CA, USA) targeted for 3x coverage.

### Whole-exome sequencing of cfDNA

WES libraries were prepared with the Accel-NGS 2S Hyb DNA Library Kit (Swift Biosciences, Ann Arbor, MI, USA) according to the manufacturer’s recommendations for SureSelect protocols. In brief, truncated adapters of the XT Compatibility Module (Swift Biosciences, Ann Arbor, MI, USA) were used in the ligation I step. The SureSelectXT Target Enrichment System for Illumina Paired-End Multiplexed Sequencing Library protocol was used to capture SureSelectXT Human All Exon, V7 (Agilent, Santa Clara, CA, USA). SureSelectXT libraries were labeled with uniquely indexed adapters in the posthybridization capture amplification step with primers from Agilent. Library quality was again assessed with a Qubit dsDNA HS Assay Kit (Thermo Fisher Scientific, Waltham, MA, USA) and TapeStation (Agilent, Santa Clara, CA, USA). Libraries were pooled in equimolar amounts and sequenced on a HiSeq 4000 paired-end 100 bp (Illumina, San Diego, CA, USA), with 300x coverage.

### Panel sequencing of cfDNA

Capture-based panel sequencing was performed via a custom brain tumor panel (Agilent Technologies, Santa Clara, CA, USA) covering the entire coding and selected intronic and promoter regions of 130 genes (0.9 Mb) of particular relevance in CNS tumors^22^. cfDNA was used without fragmentation, and the input was adjusted to 4 ng per library. Libraries were ligated with single-indexing adapters and unique molecular identifiers (UMIs; Swift Biosciences, Ann Arbor, MI, USA). Depending on the library yield, 12-16 amplification cycles were performed. Libraries were quantified via the Qubit dsDNA HS Assay Kit (Thermo Fisher Scientific, Waltham, MA, USA), and the fragment size distribution was assessed via the Bioanalyzer High Sensitivity DNA Kit (Agilent, Santa Clara, CA, USA). Libraries were pooled in equimolar amounts and sequenced at 100 bp paired-end on a HiSeq 4000 platform (Illumina, San Diego, CA, USA) targeted for 10.000x coverage.

### CNV analysis of low-coverage whole-genome sequencing data

All NGS data were transferred to the DKFZ Omics IT and Data Management Core Facility (ODCF). In-house bioinformatics workflows for sequence alignment and somatic variant calling [github.com/DKFZ-ODCF/] were performed through the One Touch Pipeline (OTP) service^41^. Briefly, sequencing reads were aligned to the human reference genome GRCh37 (hg19) ^42^ plus PhiX contig via BWA-MEM (v0.7.15)^43^. Sambamba (v0.6.5) ^44^ was used for sorting, and the alignment results were indexed. Unique molecular index (UMI) processing was performed via fgbio (v1.1.0; Fulcrum Genomics) and the Picard toolkit to collapse BAM files and identify high-quality, deduplicated molecular consensus reads that were subjected to a second round of sequence alignment with BWA-MEM. The markduplication process was not performed because of the use of UMIs. Picard CollectWgsMetrics and CollectInsertSizeMetrics (v2.21.8)^45^ estimated the sequencing quality matrix, including average sequencing coverage and distribution of template size. The CNV calling, segmentation and tumor fraction estimation tool ichorCNA (v0.3.2)^19^ was used to calculate normalized log2 ratios from read count information for each 1 MB genomic window. The Panel-of-Normal (PoN) function was used to control for the nature of cfDNA by including healthy control cfDNA samples. Integrating PoN reduced noise and corrected systematic biases arising from different sequencing platforms, sample preparation protocols, and cfDNA-specific genomic features. We implemented a protocol-specific PoN for the Accel-NGS 2S Hyb DNA Library Kit (Swift Biosciences, Ann Arbor, MI, USA). To create a PoN for samples from the Accel-NGS library, we selected UMI-deduplicated BAM files with a coverage between 0.24 and 2.35 and implemented the function “diagnose control group” from the R package NIPTeR ^46^ to create a NIPTControlGroup where samples with z-transformed chromosomal coverage stay in the range [-3,3]. This process involves screening cfDNA samples with high tumor content in cfDNA with large copy number alterations. Since the majority of cfDNA samples contained low amounts of tumor-derived cfDNA, the ichorCNA default settings were modified to improve parameter estimation. The parameters were changed as follows: –ploidy “c(2,3)” –normal “c(0.8,0.9,0.95,0.99,0.995)” –maxCN 4 –includeHOMD FALSE – estimateScPrevalence FALSE –scStates “c()” –chrTrain “c(1:22)”. LcWGS data with coverage below 0.1 were excluded from the analysis.

To facilitate comparisons between tumor and cfDNA CNVs, reported CNV events were adjusted according to the reported tumor ploidy. IchorCNA detected CNVs on the basis of sequencing coverage segmentation per 1 MB of genomic nonoverlapping windows. On the basis of the segmentation result, the software detected copy number aberrations by fitting a model with a range of parameters for tumor fractions (ichorTF) and tumor ploidy. The CNV profile with the highest likelihood score was considered. Concurrent tumor and cfDNA CNVs were called when the overlapping cfDNA segment reported the same CNV event (amplification or deletion, regardless of reported copy number) as the tumor. Moreover, the overlapping cfDNA CNV had to be covered by more than 20% of the tumor segment.

### Genome-wide methylation array data for CNV and methylation analysis

Genome-wide DNA methylation data from tumor biopsies were generated as part of the standard INFORM pipeline via the Infinium HumanMethylation450 or MethylationEPIC BeadChip (Illumina)^47^. The Conumee package (Bioconductor) was used to infer genome-wide CNVs^48^. We utilized the ‘noob’ algorithm from the minfi package in R for robust preprocessing of methylation data obtained from tumor and liquid biopsy samples. For our analysis, we specifically examined CpGs within the H19 locus, which were selected on the basis of a study by Coorens *et al*.^25^. To visualize the methylation distribution across samples, we employed the ‘densityPlot’ function available in the minfi package.

### Unique molecular indexing (UMI)

Unique molecular indexing (UMI) strategies have been shown to improve rare mutation calling in liquid biopsies. The increased duplication rates due to the high number of PCR amplification cycles to overcome cfDNA input limitations were reduced by the implementation of UMIs in lcWGS and the panel sequencing protocol (Figure S2F-G, S4E-F). The greater amount of input material used for the panel libraries (>4 ng) than for the lcWGS (100 pg) libraries resulted in fewer duplicated fragments. The UMI deduplication workflow increased the average coverage by 7% for lcWGS and the on-target coverage of panel sequencing by threefold. The increase in coverage was positively correlated with the MarkDuplication rate according to both lcWGS (Figure S2G) and panel sequencing (Figure S4F).

### In silico size selection and fragment length analysis

Paired-end sequencing reads allowed inference of the fragment lengths of cfDNA templates via the genomic locations of both ends after alignment to the reference genome. Samtools^49^ was used to select paired reads with fragment lengths between 50 and 150 bases. These size-selected samples were used to call copy-number alterations via ichorCNA, where the PoN was created from this size-selection process.

An R package, cfdnakit, was developed to employ fragment length information from lcWGS data and was published as a Bioconductor package [https://bioconductor.org/packages/cfdnakit.html]. This package includes a set of functions to extract read information from a BAM file. Briftly, reads overlapping DUKE and DAC blacklisted regions^50^ and (the) centromere(s) were excluded from downstream analysis. The short–fragment ratio (SLRatio) of a sample is defined as the ratio of fragments from 100--150 nt to fragments 151-250 nt in length.

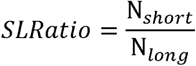

where N_*short*_ is the number of short fragments and where N_*long*_ is the number of long fragments. In addition, the number of short and long reads was calculated per 1 Mb nonoverlapping genomic region. For each bin, the read counts of both short and long fragments were corrected to account for percent GC content and mappability via the LOESS regression model. SLRatio per bin was then calculated on the basis of the corrected read count. We normalized bin SLRatio by their median genome-wide SLRatio, noted as the z score. For each bin, z scores in test samples were normalized by the z score of the same bin from process-matched PoN samples. Finally, bins were segmented via circular binary segmentation (CBS)^51,^ and the median z score per segment (Z_*segment*_) was calculated. The CtDNA estimation score (CES) is defined as follows:

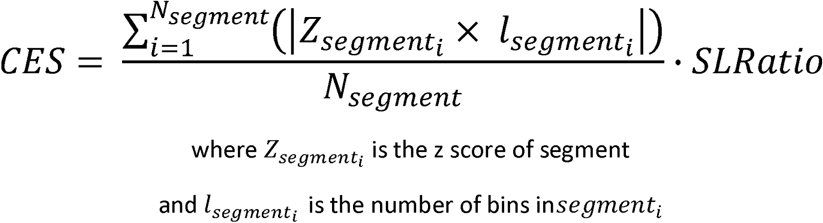

### Whole-exome sequencing analysis

Similar to WGS analysis, the OTP service was used to align WES sequencing data and call somatic variants. Picard CollectWgsMetrics, CollectOxoGMetrics, and CollectInsertSizeMetrics estimate the average sequencing coverage, the error rate from the oxidation of guanine to 8-oxoguanine, and the distribution of template sizes according to on-target reads. The samples were subjected to quality thresholds of coverage at 60x, and an average Phred score of 30 was calculated from the substitutions C(C>A or G>T). Somatic SNV and indel calling from matching tumor/control WES were processed via the ODCF in-house SNVCallingWorkflow and IndelCallingWorkflow through the OTP service. The minimum confidence score was set to 8. Nonsynonymous somatic mutations were selected for downstream analysis. The same workflows were also used for somatic SNV and indel calling from matching cfDNA and healthy controls with the following options: -t 500 -c 0 -x 1 -l 1 -e 0 to allow detection with a low allelic fraction. Both the SNV and indel calling workflows are available at [github.com/DKFZ-ODCF/]. We used the tumor VCF file containing high-confidence nonsynonymous somatic variants as a reference to interrogate variant abundance in cfDNA from patients. An in-house bash script was used to annotate the existence of a tumor variant allele by looking up read pileup information from cfDNA. We considered read pileup information with a minimum read mapping quality of 1 and a base quality of 20. Tumor variants were reported to be present in the cfDNA sample if at least one read supported the tumor allele and if at least 11 reads covered the affected genomic region. We used PureCN^52^ for CNV calling on WES data. To create a PoN, we selected cfDNA WES samples with on-target coverage between 142 and 269. Subsequently, cfDNA samples from patients with fewer than three tumor variants were included. BAM files of selected samples were processed by GATK Mutect2 in tumor-only mode for calling both germline and somatic variants with the setting --max-mnp-distance 0 --min-base-quality-score 20 --annotation BaseQuality --read-filter MappingQualityReadFilter --read-filter OverclippedReadFilter --minimum-mapping-quality 30 --read-filter FragmentLengthReadFilter --min-fragment-length 30. After that, we combined VCF files with GATK CombineVariants with the option --minimumN 3. This setting retained only variants commonly found in at least three samples. Finally, PureCN CNV calling was applied to cfDNA WES samples with the following settings to (also) allow the detection of CNV in samples with lower tumor purity: --minpurity 0.05 --minaf 0.01 --error 0.0005 --maxploidy 3 --maxcopynumber 8 --padding 25 --model betabin --funsegmentation PSCBS –postoptimize.

### Panel sequencing analysis

Sequence alignment was performed via the ODCF in-house bioinformatics workflow. The workflow aligns sequencing reads to the human reference genome GRCh37 (hg19) plus the PhiX contig. As described in the WGS section, fgbio (v1.1.0; Fulcrum Genomics) and Picard were used to manipulate BAM files and call molecular consensus reads from UMI information. Picard SamToFastq extracted consensus reads into FASTQ format. These consensus reads were aligned to the reference genome again by using BWA-MEM without marking duplicate reads. Picard CollectWgsMetrics and CollectOxoGMetrics (v2.21.8) were used to estimate the sequencing quality, including the average sequencing coverage and level of pre-PCR artifacts from the oxidation of guanine to 8-oxoguanine of on-target reads. For WES, tumor nonsynonymous somatic variants were used as a reference to look for concordance with the cfDNA of patients. Read pileup information from reads with a minimum mapping quality of 1 and a minimum base quality of 20 was included. Tumor variants were considered to be represented in cfDNA if at least one read supported the tumor allele at a minimal coverage of 11.

## Data Availability

The high-throughput sequencing datasets generated for this study can be found in EGA European Genome-Phenome Archive under accession number EGAS50000000393.

## Data Visualization and Statistical Analysis

Patient data management was performed via GraphPad Prism 8 (GraphPad, Software, Inc., La Jolla, CA, USA). Cohort representation diagrams and tumor-informed oncoplots were visualized via ComplexHeatmap. Nonparametric comparisons between groups were performed with the Wilcoxon rank-sum test and the Kolmogorov–Smirnov test in the R environment (v4.0.0). P values of <0.05 were considered statistically significant, with p values represented as follows: * p < 0.05, ** p < 0.01, *** p < 0.001, and **** p < 0.0001. “ns” denotes differences in means that were not significant. All error bars shown represent the standard deviation unless otherwise stated. Graphical illustrations of patient disease courses were created with BioRender.com.

## Author’s contribution

**K.K. Maass:** Methodology, data curation, formal analysis, validation, investigation, visualization, conceptualization, writing–original draft, writing–review and editing. **P. Puranachot:** Data curation, software, validation, writing–review and editing. **P.S. Schad:** Investigation, data curation, visualization, validation, writing–review and editing. **A.M.E. Finster:** Investigation, writing–review and editing. **S. Volz:** Investigation, data curation, writing–review and editing. **T.T. Fischer:** Investigation, writing–review and editing. **B.C. Jones:** Data curation, investigation, writing–review and editing. **K. Schramm:** Investigation, writing–review and editing. **S.C. Henneken:** Investigation, writing–review and editing. **N. Simon:** Investigation, writing–review and editing. **S. Montigel:** Investigation, writing–review and editing. **T. Wedig:** Investigation, writing–review and editing. **N. Schwarz:** Investigation, writing–review and editing. **C. Zuliani:** Data curation, writing–review and editing. **P. Fiesel:** Investigation, writing–review and editing. **C. Previti:** Formal analysis, validation, investigation, visualization, methodology, writing–review and editing. **G. Balasubramanian**: Investigation, writing–review and editing. **F. Iser:** Investigation, writing–review and editing. **J. Meyer:** Investigation, writing–review and editing. **C.M. van Tilburg:** Investigation, writing–review and editing. **T. Milde:** Investigation, writing–review and editing. **O. Witt:** Investigation, writing–review and editing. **C. Rossi:** Investigation, writing–review and editing. **M. Sparber-Sauer:** Investigation, writing–review and editing. **S. Zimmermann:** Investigation, writing–review and editing. **T. Lehrnbecher:** Writing–review and editing. **M. Lauten:** Investigation, writing–review and editing. **M. Sill:** Formal analysis, visualization, methodology, writing–review and editing. **N. Jäger:** Investigation, writing–review and editing. **R.J. Autry:** Formal analysis, visualization, methodology, writing–review and editing. **P.A. Northcott:** Writing–review and editing. **F. Sahm:** Investigation, writing–review and editing. **D.T.W. Jones:** Conceptualization, resources, writing–review and editing. **S.M. Pfister:** Conceptualization, supervision, investigation, writing–review and editing. **B. Brors:** Conceptualization, resources, supervision, funding acquisition, investigation, methodology, writing–review and editing. **K.W. Pajtler:** Conceptualization, resources, supervision, funding acquisition, investigation, methodology, writing–review and editing.

## Acknowledgement

We would like to convey our heartfelt thanks to Carsten Maus, Erjia Wang (Genomics and Proteomics Core Facility, DKFZ). Lena Weiser, Gregor Warsow (Omics IT and Data Management Core Facility, DKFZ) for their highly dedicated support in data management and processing. We thank Ute Ernst, Franziska Petermann, Angela Schulz, and Stephan Wolff from the NGS Core Facility, German Cancer Research Center (DKFZ), for providing excellent consultancy regarding low-input sequencing library adaptations, capture efficiencies, and sequencing modalities. The authors express their sincerest gratitude to all patients, families, clinical care teams, operations staff, and research personnel from participating institutions for their contributions.

The INFORM program is financially supported by the German Cancer Research Center (DKFZ), several German health insurance companies, the German Cancer Consortium (DKTK), the German Federal Ministry of Education and Research (BMBF), the German Federal Ministry of Health (BMG), the Ministry of Science, Research and the Arts of the State of Baden-Württemberg (MWK BW); the German Cancer Aid (DKH), the German Childhood Cancer Foundation (DKS), RTL television, the aid organization BILD hilft e.V. (Ein Herz für Kinder) and the generous private donation of the Scheu family.

---

**Supplementary Figure 1.**
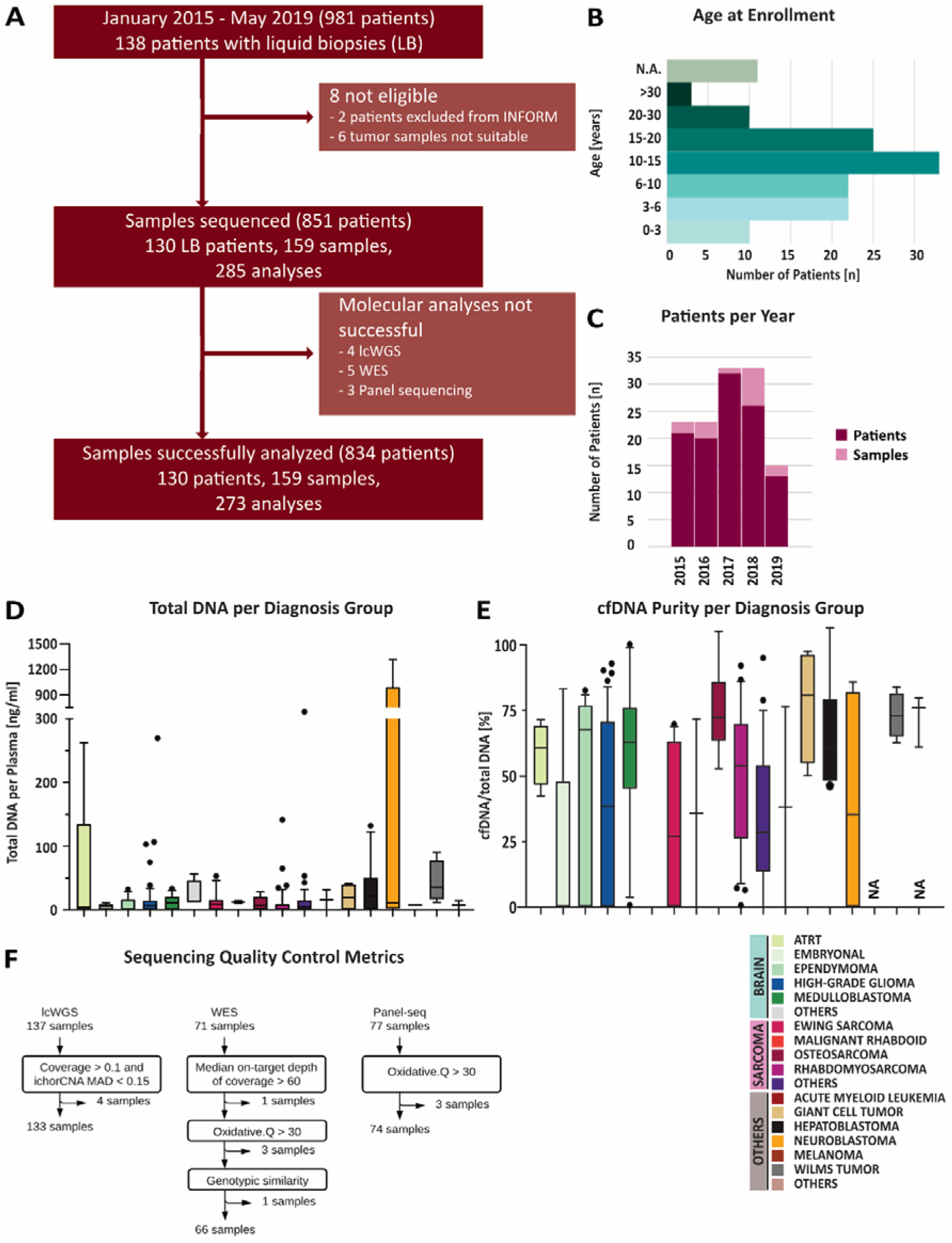
Cohort Characteristics and Quality Metrics. **A)** Cohort overview of INFORM patients registered between January 2015 and May 2019 with liquid biopsy blood plasma material available. **B)** Age distribution in years of cohort patients. **C)** INFORM patient registrations with available liquid biopsy material over time (dark red - patients, light red - liquid biopsy samples). **D)** Total amounts of DNA isolated per ml plasma of the indicated diagnosis groups. **E)** Purity of isolated cell-free DNA (cfDNA) calculated as the ratio of cfDNA to total DNA in % determined by Bioanalyzer fragment length measurements. **F)** Quality control metrics for low-coverage whole-genome sequencing (lcWGS), whole-exome sequencing (WES) and panel sequencing data revealing the final liquid biopsy cohort for orthogonal sequencing comparison of 159 samples being subjected to 273 analyses. Of all 137 lcWGS samples, 4 samples were excluded due to genomic coverage of lower than 0.1x or ichorCNA MAD less than 0.15. Of all 71 WES samples, 1 sample was excluded due to an on-target coverage lower than 60x and 3 samples were excluded due to an excessive level of oxidative artefacts. Of all 77 panel sequencing samples, 3 had to be excluded due to high oxidative artefact rates. MAD – median absolute deviation as QC output of ichorCNA.

**Supplementary Figure 2.**
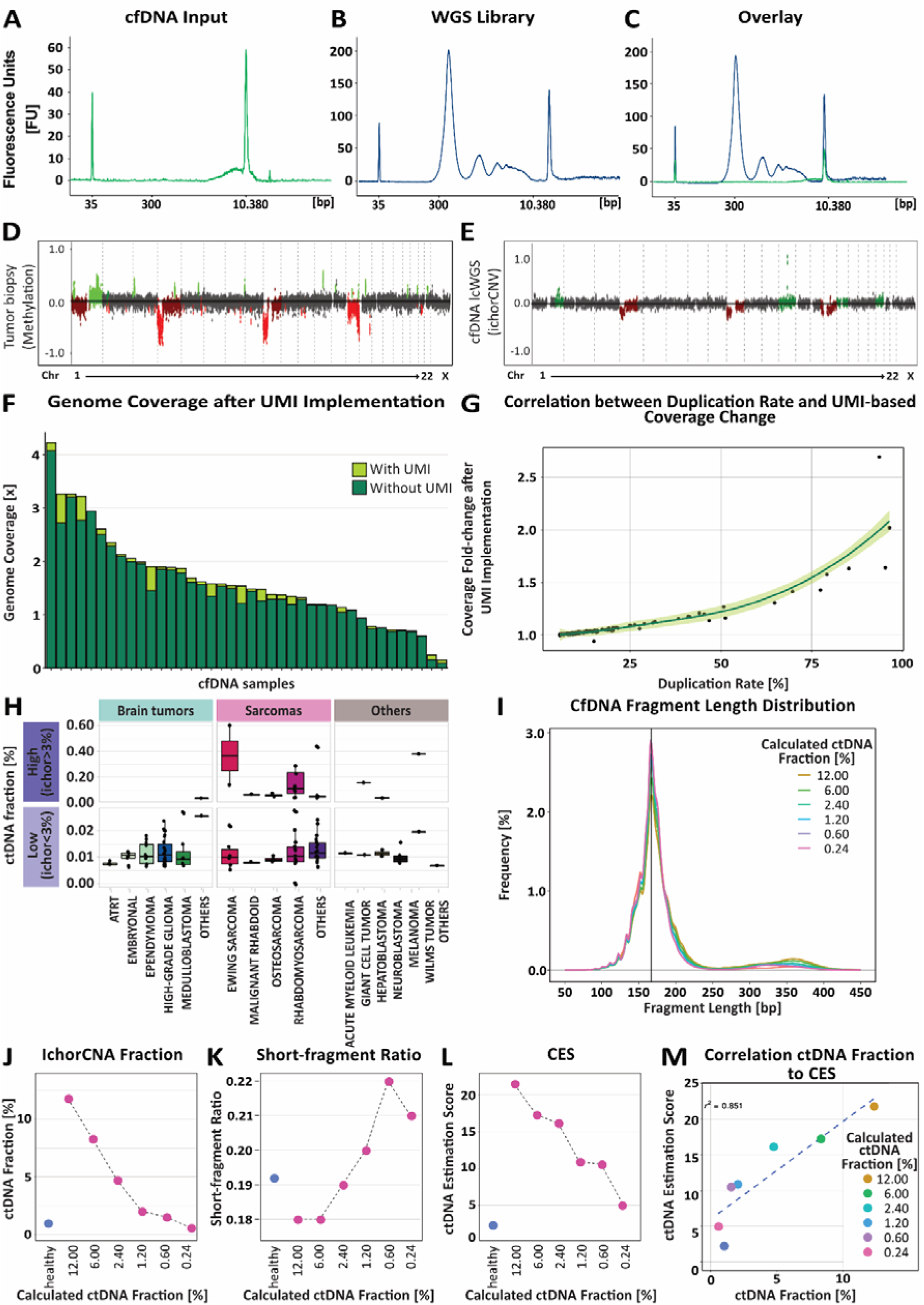
Increased Genome Coverage of Low-coverage Whole-Genome Sequencing (lcWGS) through Implementation of Unique Molecular Identifier (UMI) **A)** Bioanalyzer size profile for cell-free DNA (cfDNA) isolated from plasma of a patient with alveolar rhabdomyosarcoma and **B)** the respective library for lcWGS. **C)** Overlay plot of cfDNA and library size profiles illustrates enrichment of DNA by a library protocol adapted for low-input material. **D)** EPIC array-derived copy number variation (CNV) profile of tumor DNA and **E)** corresponding lcWGS-based CNV profile (library shown in **B)** from plasma cfDNA for patient in A. This exemplary case illustrates the feasibility of lcWGS-based CNV detection from minimal cfDNA inputs. **F)** Implementation of UMI-based deduplication (light green) improved genome coverage over MarkDuplication (dark green) by on average 7% in lcWGS samples. **G)** Increasing coverage in lcWGS showed positive correlation with the degree of duplicated reads removed by MarkDuplication. **H)** Calculated ctDNA fractions for indicated tumor entities plotted for high (ichor>3%, dark purple) and low (ichor<3%, light purple) ctDNA fraction groups. Data revealed high ctDNA fractions for Ewing sarcomas, rhabdomyosarcomas, and other sarcomas as well as for Wilms tumors while brain tumors and other solid tumors mostly showed low ctDNA fractions. **I)** *In silico* fragment-length distribution of a titration series using cfDNA from a Rhabdomyosarcoma that was spiked into healthy control cfDNA at indicated ratios (ichorCNA ctDNA fraction 12%, 6%, 2.4%, 1.2%, 0.6% and 0.24%). Graph shows that short fragments increase with higher dilution steps. **J)** Course of ichorCNA-based ctDNA fraction, **K)** short-fragment ratio, and **L)** ctDNA estimation score (CES) with increasing dilution as indicated in **I)**. **M)** Correlation of ctDNA fractions to the CES for the dilution series resulted in a correlation coefficient of 0.85.

**Supplementary Figure 3.**
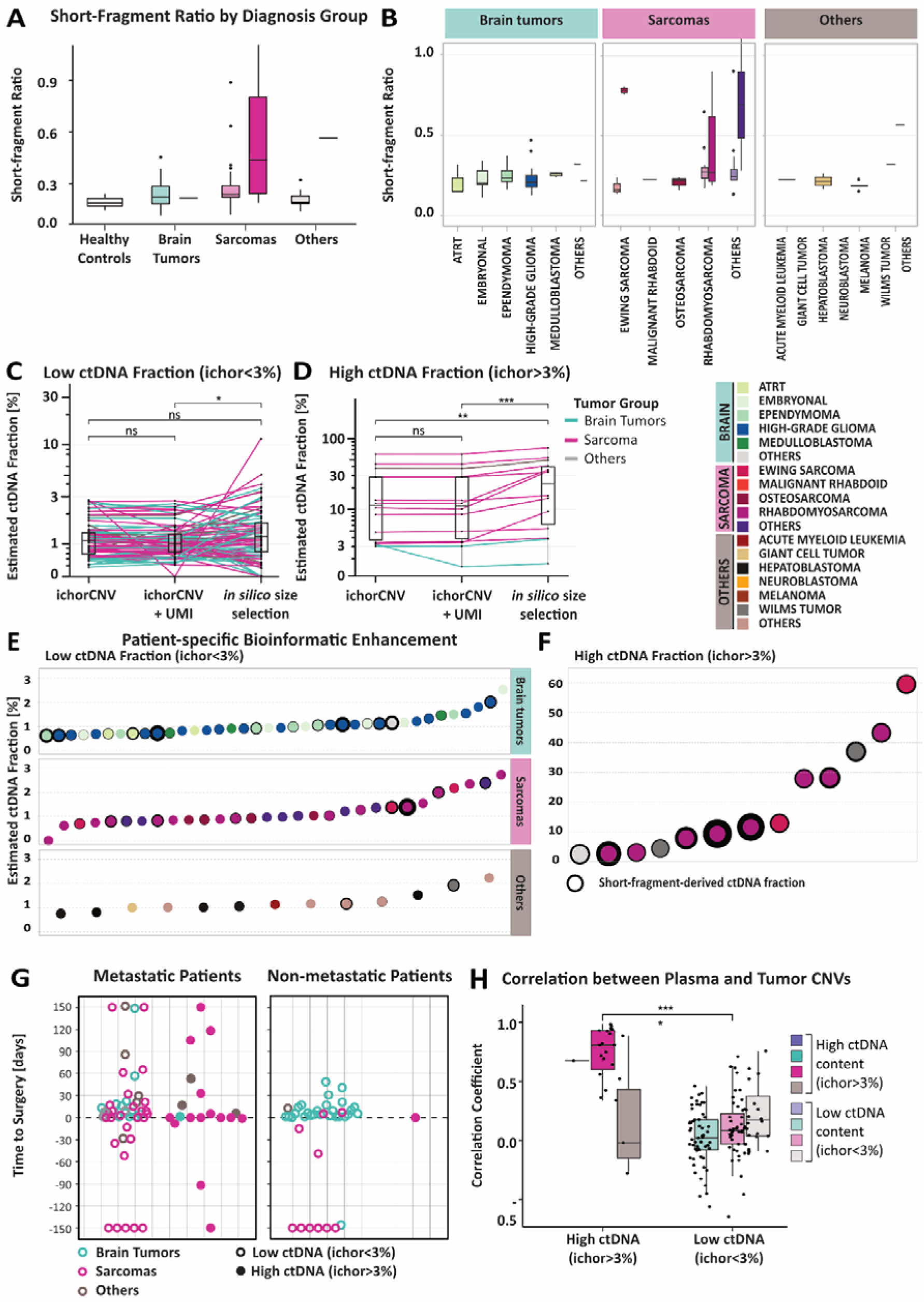
Low-Coverage Whole-Genome Sequencing (LcWGS) of Liquid Biopsies Recapitulated Tumor Copy Number Variations (CNVs) with High Sensitivity and Specificity. **A)** Box plot representation of short-fragment ratio in plasma samples of healthy controls, brain tumor, sarcoma, and patients with other tumors showing strong enrichment of short cell-free DNA (cfDNA) fragments in sarcoma patients. **B)** Distribution of short-fragment ratios for indicated tumor entities revealing high enrichment of short fragments in Ewing sarcomas, Rhabdomyosarcomas and other sarcomas. **C-D)** Estimated circulating tumor DNA (ctDNA) fraction based on ichorCNA for **C)** low (ichor<3%) and **D)** high (ichor> 3%) ctDNA samples. Plasma ctDNA fraction was slightly improved bioinformatically by implementing unique molecular identifier (UMIs) but significant higher ctDNA fractions could be extracted by performing *in silico* short fragment size selection. P-values were calculated by paired Wilcoxon signed rank test with *= p<0.05** = p<0.01, *** = p<0.001. **E-F)** Increased coverage by UMI implementation resulted in improved ctDNA fraction detection shown for individual patients. **E)** Tumor detection in plasma samples of low ctDNA patient group (ichor<3% ctDNA fraction) revealed strongest improvement for brain tumors (upper graph), followed by sarcomas (middle graph), and other tumors. IchorCNA estimated ctDNA fraction is plotted on the y-axis and the increase of the estimated ctDNA fraction following *in silico* size selection is indicated by black circles drawn to scale. **F)** Tumor detection in plasma samples of high ctDNA patient group (≥3% ctDNA fraction) revealed stronger improvement in ctDNA fraction estimation for sarcomas over other tumors. Tumor entities are indicated with color code shown in legend above. **G)** Time point of plasma sampling is illustrated as days prior (-) or post (+) surgery for patients with or without clinically apparent metastases. cfDNA samples are labeled according to high (ichor >3%; filled circles) and low (ichor <3%; empty circles) ctDNA fractions. **H)** Pearson correlation coefficient of genomic log2 ratio as surrogate marker for copy number variation (CNV) similarities between tumor and matched cfDNA samples showed higher correlation for high (ichor>3%) than for low (ichor<3%) ctDNA groups irrespective of diagnosis groups.

**Supplementary Figure 4.**
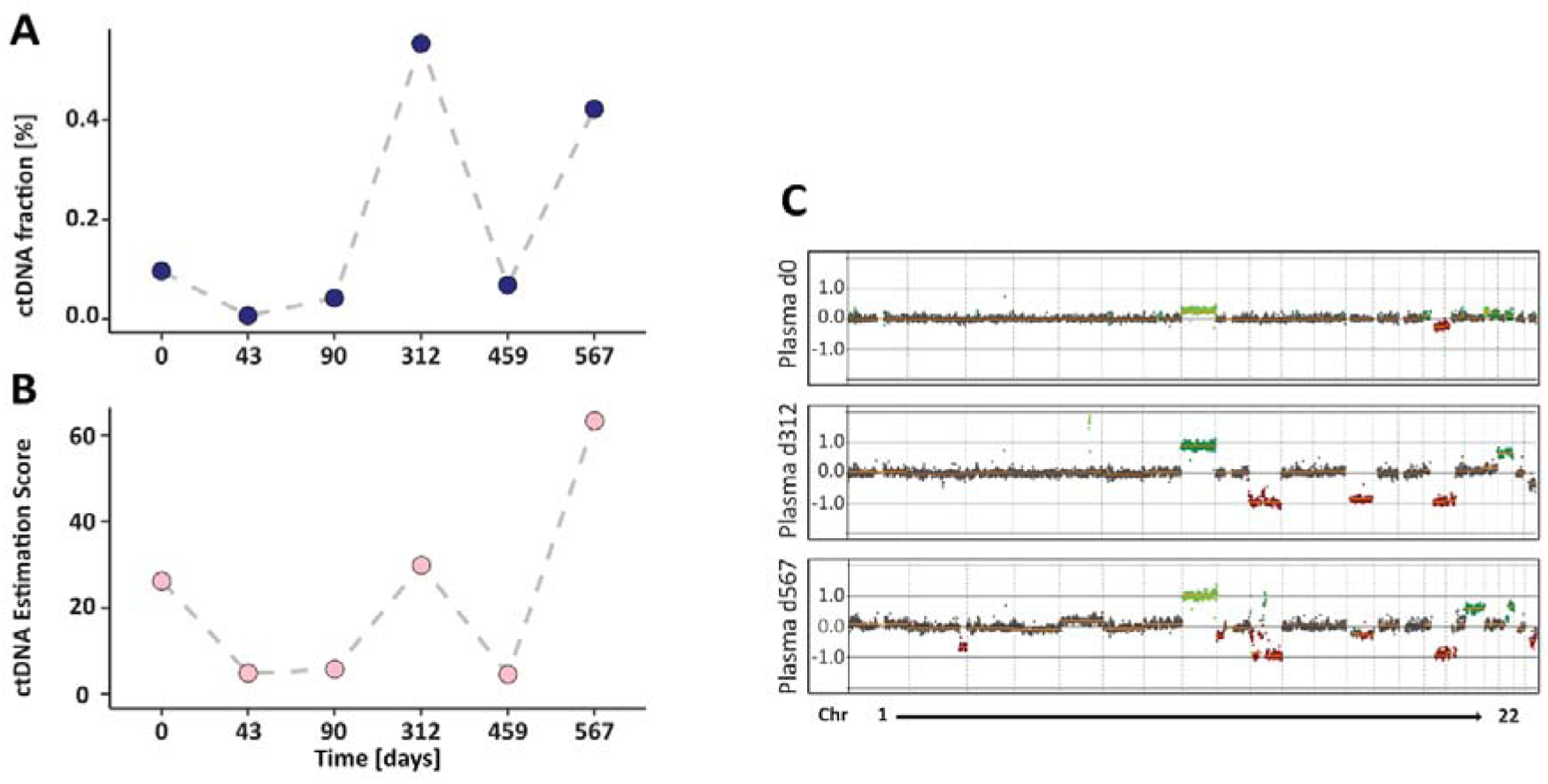
CES Validation in Independent Ewing Sarcoma Cohort^21^. **A)** IchorCNA estimated ctDNA fraction and **B)** ctDNA Estimation Score (CES) plotted over clinical course of Ewing Sarcoma patient. Clinical events include diagnosis (day 0), first relapse (day 312), second relapse (day 567), and death (day 654) are annotated. Both metrics increase in concordance with relapse events, supporting their utility as non-invasive biomarkers for disease burden and progression. **C)** Genome-wide CNV profiles derived from cfDNA WGS are depicted at diagnosis (day 0), first relapse (day 312), and second relapse (day 567).

**Supplementary Figure 5.**
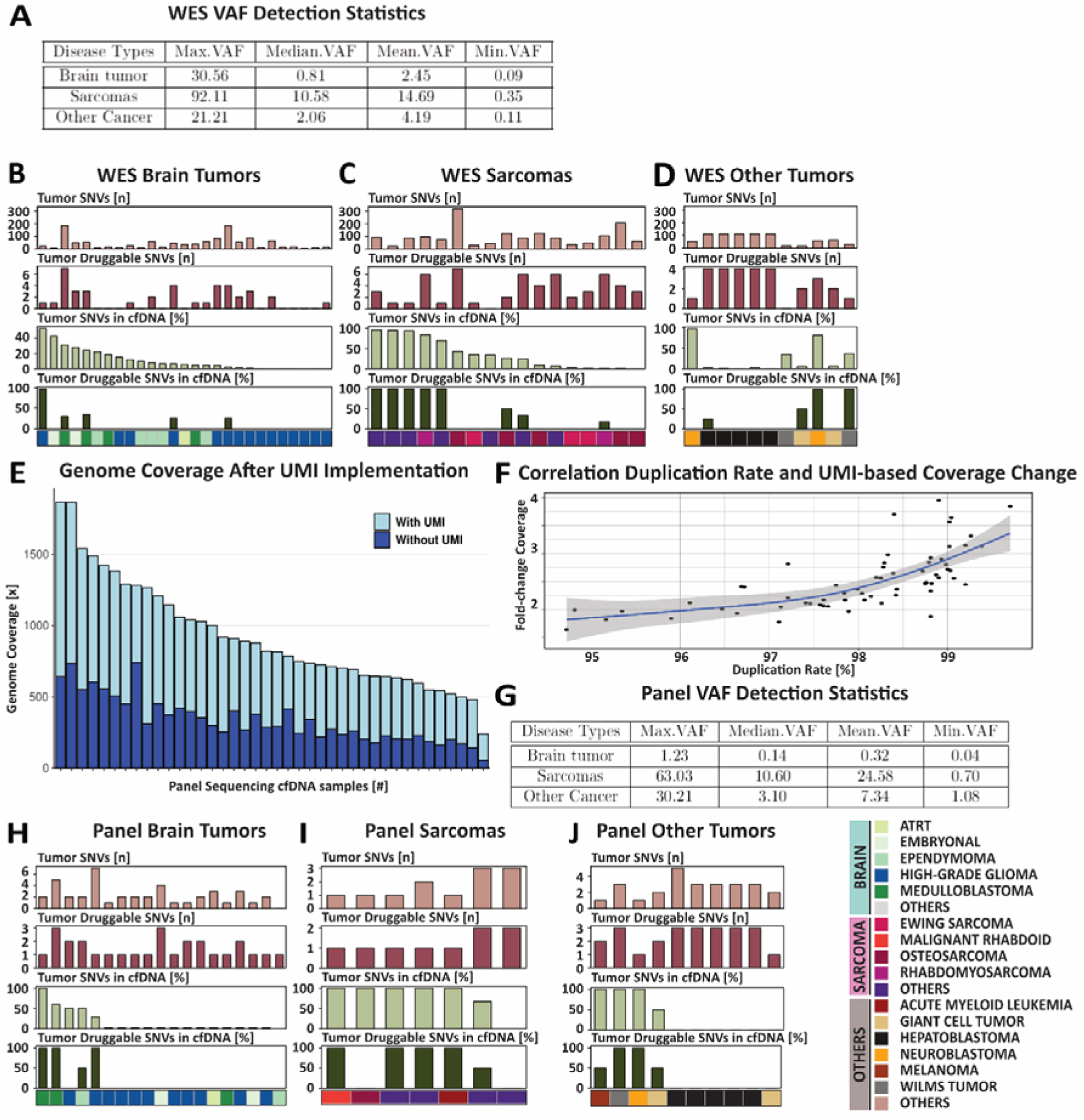
Sequencing Specifics and Single Nucleotide Variant (SNV) Detection Rate of Whole-Exome Sequencing (WES) and Targeted Panel Sequencing. **A)** Descriptive variant allele frequency (VAF, %) of tumor-informed SNVs detected in cell-free DNA (cfDNA) WES data. B-D) Bar plots representation for patient-specific absolute number of tumor SNVs [n], tumor druggable SNVs [n], relative detection of tumor SNVs in cfDNA [%], and relative detection of tumor druggable SNVs in cfDNA [%] WES data **B)** of individual brain tumor, **C)** sarcoma, and **D)** other tumor liquid biopsies. **E)** On-target coverage of cfDNA panel sequencing data compared between regular MarkDuplication procedure and the UMI-based deduplication workflow. UMI-based deduplication (light blue) improved on-target coverage by around 3-fold to regular MarkDuplication (dark blue). **F)** Increasing on-target coverage in panel sequencing data correlated with high degree of duplicated reads removed by MarkDuplication procedure. **G)** Descriptive variant allele frequency (VAF, %) of tumor-informed SNVs detected in cfDNA panel sequencing data. H-J) Bar plots of panel sequencing data of individual brain tumor, **I)** sarcoma, and **J)** other tumor patients.

**Supplementary Figure 6.**
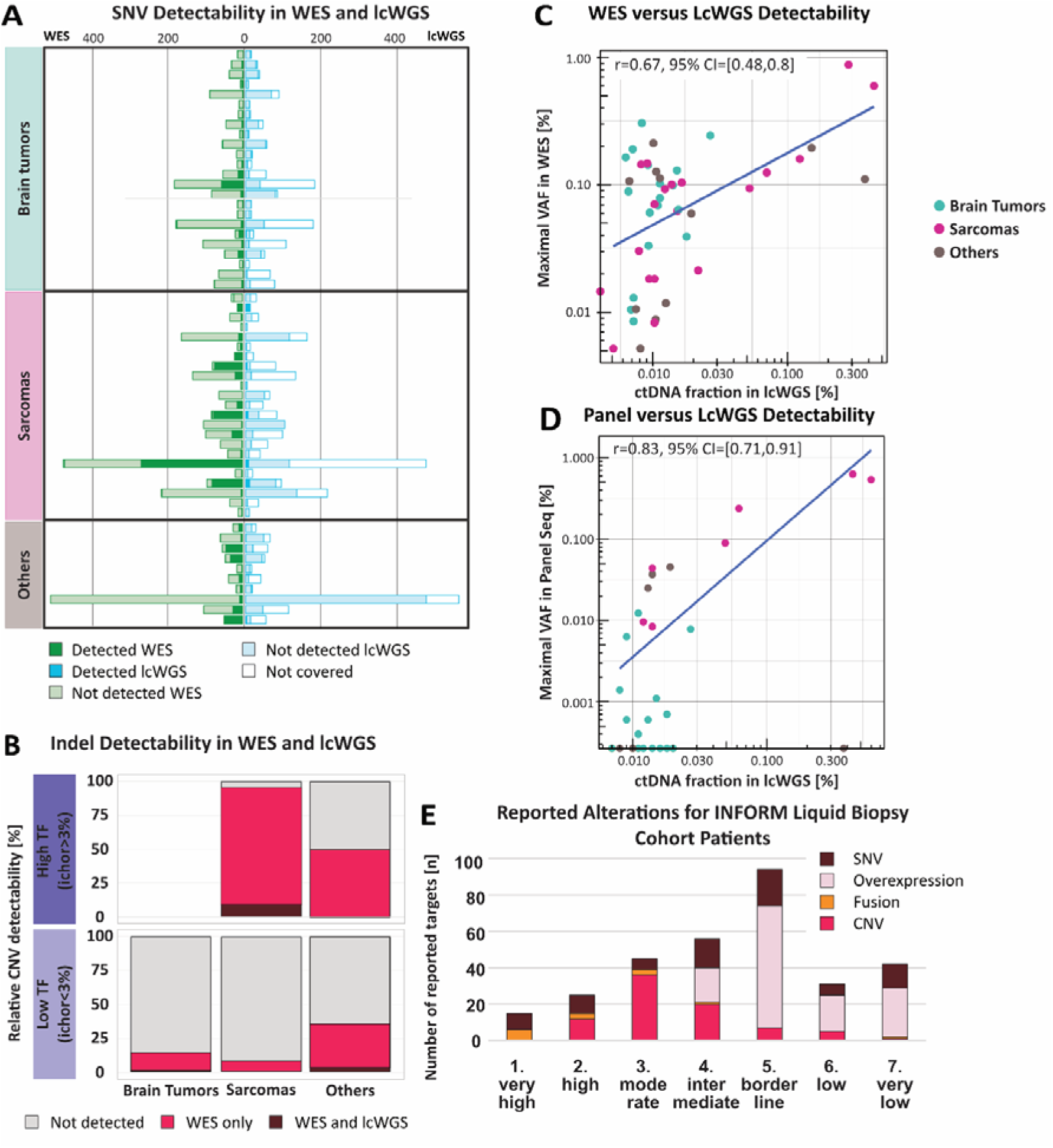
Comparison of Orthogonal NGS Approaches for Detection of Tumor-derived Alterations in cell-free DNA (cfDNA) **A)** Plasma-based detection of single nucleotide variants (SNVs) that were reported for matched tumors applying whole-exome (WES) and low-coverage whole-genome sequencing (lcWGS). WES: tumor SNVs detected (green) or not detected (light green) in cfDNA despite sufficient coverage. LcWGS: tumor SNVs detected (blue) or not detected (light blue) in cfDNA despite sufficient coverage. Tumor SNVs for which no sufficient coverage was reached are indicated with white bars. **B)** Detection of small insertions and deletions (Indels) in cfDNA applying WES and lcWGS. Red – Indel detected by WES only, dark red – Indel detected by WES and lcWGS, grey – not detected by any method. **C)** Correlation of lcWGS-derived cell-free tumor DNA (ctDNA) fractions [%] with maximal variant allele frequencies (VAF) of detected tumor-informed mutations in WES [%] and **D)** in panel sequencing [%]. **E)** Overview of alteration types and target priority levels reported for corresponding tumors of patients in the liquid biopsy cohort within the INFORM molecular tumor board. Dark red – SNVs, light pink – overexpression, orange – fusions, red – copy-number variations (CNVs).

**Supplementary Figure 7.**
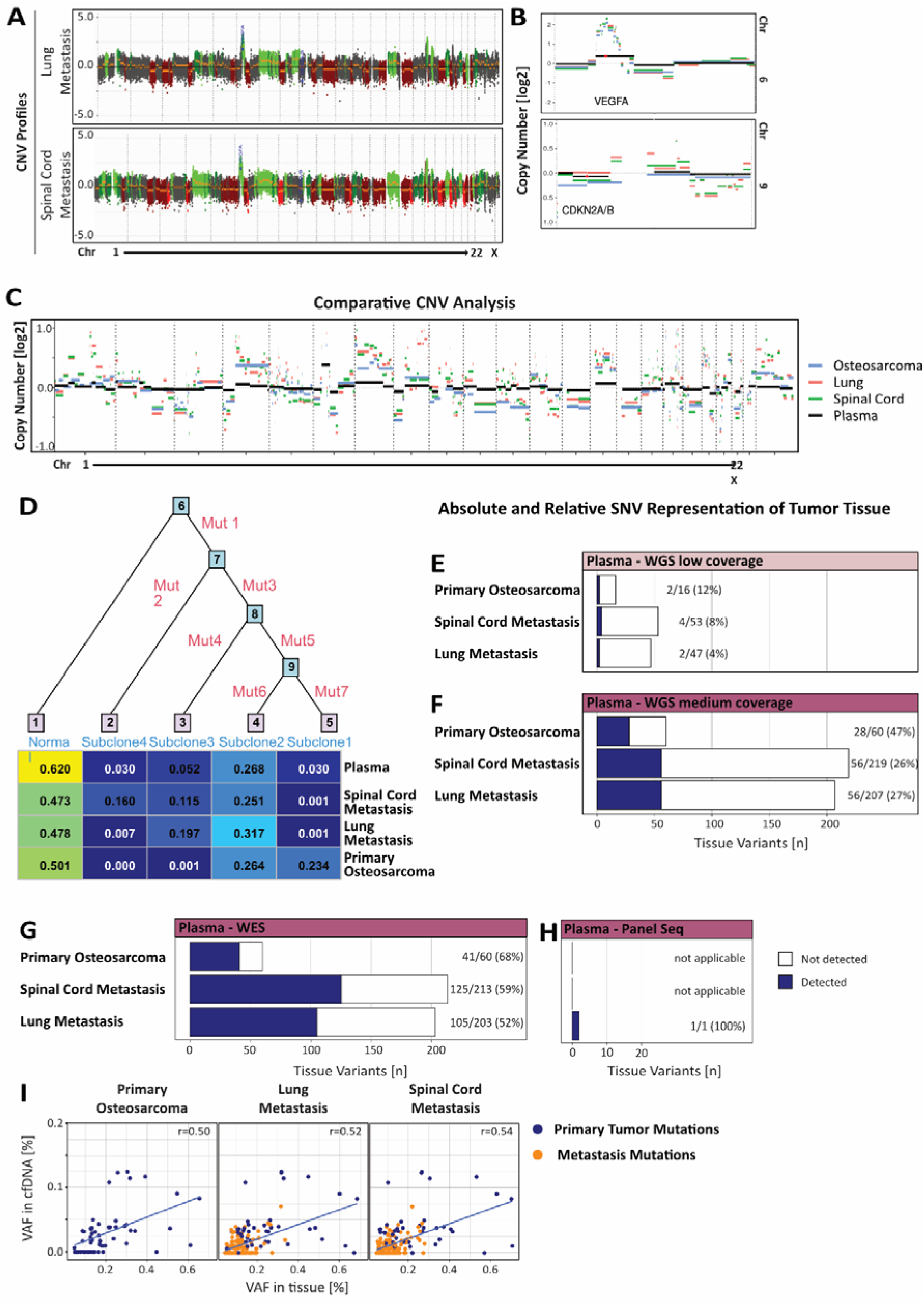
Liquid Biopsy-based Detection of Temporal Tumor Heterogeneity. **A)** Single nucleotide variant (SNV)-based phylogenetic trees depict variable clonal diversification. Branch lengths correspond to the total numbers of SNVs. Relative contribution of indicated subclones per tumor site and in the plasma sample. **B)** Individual CNV profiles generated from whole-exome sequencing (WES) data from tumor tissue of lung and spinal cord metastases. **C)** Comparative CNV analysis depicted as overlay plot of the primary osteosarcoma (blue), a lung mtastasis (41 months after diagnosis, green), a spinal cord metastasis (52 months after diagnosis, red) and plasma cfDNA (52 months after diagnosis, black). E-H) Absolute and relative detection rates of somatic tumor SNVs of primary osteosarcoma, spinal cord and lung metastases. E-F) Whole-genome sequencing (WGS) with **E)** low and **F)** medium coverage revealed higher absolute numbers of somatic mutations in cell-free DNA (cfDNA) with medium coverage, while relative numbers showed similar patterns. **G)** WES showed higher absolute and relative detectability of somatic SNVs than other sequencing methods. **H)** Panel sequencing only covered Notch1 mutation present in both the lung metastasis and cfDNA. **I)** Evolution of tumor SNVs and their respective variant allele frequencies (VAF) in tumor tissue (primary/metastasis, WES) and plasma-derived cfDNA (WES). Correlation plot of VAF in cfDNA and tissue of indicated tumor sites. Dark blue indicates SNVs detected in the primary osteosarcoma, orange indicates SNVs that evolved over time (*r=0.50 (Osteosarcoma), r=0.52 (Lung Metastasis), r=0.54 (Spinal Cord Metastasis*).

**Supplementary Figure 8.**
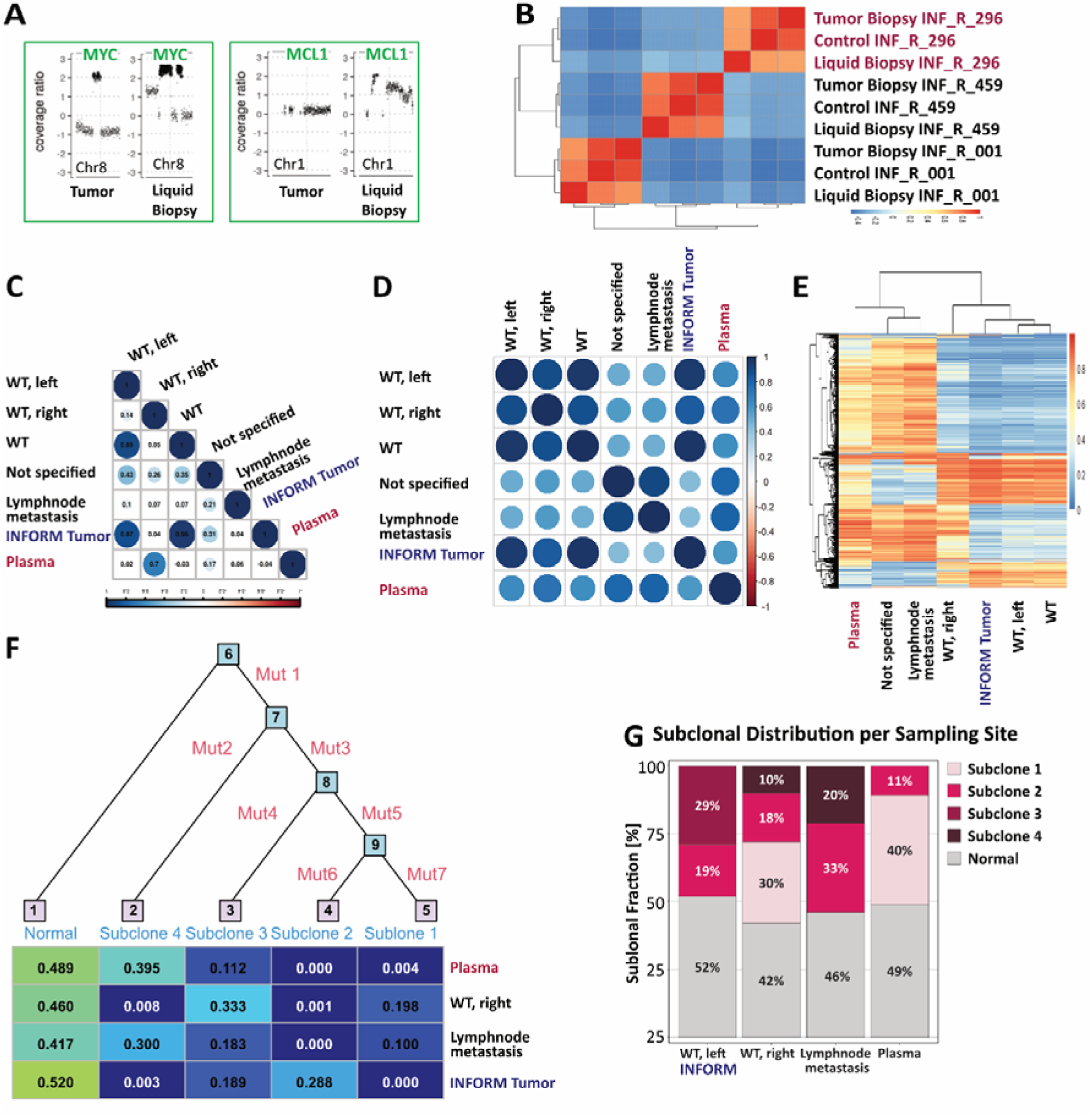
Liquid Biopsy-based Detection of Spatial Tumor Heterogeneity. **A)** MYC and MCL-1 amplifications as presented in the INFORM molecular tumor board here illustrated for tumor and liquid biopsy. **B)** Genotyping correlation matrix of 2 additional randomly selected patients including tumor biopsy, germline control and plasma cell-free DNA (cfDNA) each. The correlation coefficient suggests compatibility of all specimens from patient with Wilms tumor (WT). **C)** Pearson correlation of copy number variations (CNVs) for indicated tumor sites and plasma. Correlation coefficient indicated by color and circle size. **D)** Variance weighted Pearson correlation of methylation patterns from indicated biopsy samples revealed highest similarity between the plasma cfDNA clone and the Wilms tumor (WT) from the right site. **E)** Hierarchical clustering of the top 20.000 CpG probes with highest standard deviation indicated a methylation pattern unique to the WT of the right kidney and plasma clone. **F)** Single nucleotide variant (SNV)-based phylogenetic trees depict variable subclonal diversification. Branch lengths correspond to the total numbers of SNVs. **G)** Relative contribution of indicated subclones per tumor site and in the plasma sample

